# Associations of polygenic scores for sleep traits with cognitive function

**DOI:** 10.64898/2026.08.14.26360402

**Authors:** Xinye Qiu, Annah Wyss, Yu Zhang, Brian Spitzer, Susan Redline, Michael Brown, Xiang Li, Chloé Sarnowski, Jan Bressler, Tanika N. Kelly, Bing Yu, Alanna C. Morrison, Charles DeCarli, Qibin Qi, Robert Kaplan, Wassim Tarraf, Myriam Fornage, Joshua C. Bis, Sina A. Gharib, Jerome I. Rotter, Stephen S. Rich, Peter Y. Liu, Kent D. Taylor, Xiuqing Guo, Susan Heckbert, Alexis C. Wood, Hector M. Gonzalez, Carmen R. Isasi, Melissa Lamar, Tamar Sofer

**Affiliations:** CardioVascular Institute, Beth Israel Deaconess Medical Center, Boston, Massachusetts, USA; Department of Medicine, Harvard Medical School, Boston, Massachusetts, USA; Division of Sleep and Circadian Medicine, Brigham and Women’s Hospital, Boston, Massachusetts, USA; Human Genetics Center, Department of Epidemiology, School of Public Health, The University of Texas Health Science Center at Houston, Houston, Texas, USA; Department of Medicine, Division of Endocrinology, Diabetes and Metabolism, University of Illinois Chicago, Chicago, Illinois, USA; Department of Neurology, University of California Davis Medical Center, Sacramento, California, USA; Department of Epidemiology & Population Health, Albert Einstein College of Medicine, Bronx, New York, USA; Institute of Gerontology & Department of Healthcare Sciences, Wayne State University, Detroit, Michigan, USA; Brown Foundation Institute of Molecular Medicine, McGovern Medical School, The University of Texas Health Science Center at Houston, Houston, Texas, USA; Cardiovascular Health Research Unit, University of Washington, Seattle, Washington, USA; Division of Pulmonary, Critical Care and Sleep Medicine, University of Washington, Seattle, Washington, USA; The Institute for Translational Genomics and Population Sciences, Department of Pediatrics, The Lundquist Institute for Biomedical Innovation at Harbor-UCLA Medical Center, Torrance, California, USA; Department of Genome Sciences, School of Medicine, University of Virginia, Charlottesville, Virginia, USA; Children’s Nutrition Research Center, Baylor College of Medicine, Houston, Texas, USA; Department of Neurosciences, University of California San Diego, San Diego, California, USA; Rush Alzheimer’s Disease Center and the Department of Psychiatry and Behavioral Sciences, Rush University Medical Center, Chicago, Illinois, USA; Department of Biostatistics, Harvard T.H. Chan School of Public Health, Boston, Massachusetts, USA

**Keywords:** sleep, cognitive function, meta-analysis, long sleep, obstructive sleep apnea

## Abstract

**INTRODUCTION:** Polygenic scores (PGSs) for sleep traits are potentially more stable, and less subject to confounding than measured sleep traits. Leveraging data from five observational cohorts, we aim to assess the associations between PGS for six common sleep traits, and global cognitive function (GCF) among middle-aged to older adults.

**METHODS:** In each cohort, GCF was defined as the first principal component (PC) of multiple cognitive measures and was projected from baseline (first selected visit) to measures from a subsequent follow up visit. Poor GCF was defined as having GCF < 1 standard deviation (SD) of the age-adjusted GCF distribution median. We estimated sleep PGS associations with baseline GCF, poor baseline GCF, GCF change between baseline and follow-up, and incident poor GCF at follow-up. Models adjusted for age, sex, study center, race/ethnicity, genetic PCs, and education. Results were meta-analyzed via fixed effects meta-analysis.

**RESULTS:** Estimates are reported per 1 SD increase in PGS. A higher PGS for long sleep was associated with lower GCF at baseline (estimate = -0.02 SD, 95% CI: -0.03 to 0.00, p = 0.01) and higher risk of poor GCF at baseline (odds ratio, OR = 1.04, 95% CI: 1.00 to 1.09, p = 0.06). In addition, a higher PGS for BMI-adjusted OSA was associated with higher risk of poor GCF at baseline (OR = 1.11, 95% CI: 1.00 to 1.22, p = 0.04).

**DISCUSSION:** Genetic predisposition to long sleep and OSA is associated with poorer cognitive function in a meta-analysis of more than 20,000 middle-aged and older adults.

## 1. Background

The spectrum of cognitive aging including cognitive impairment represents an increasing public health challenge as the global population continues to age.^1^ A number of risk factors have been identified for cognitive impairment, including genetic, demographic and lifestyle measures. In particular, several studies have shown that suboptimal sleep is associated with cognitive health outcomes and changes in global cognitive function.^2–11^ For sleep duration, studies often show U-shaped risk curves with long and short sleep duration associated with poorer cognitive performance.^5,7,8,11,12^ Likewise, studies of insomnia and obstructive sleep apnea (OSA) overall suggest associations with poorer cognitive performance, though results vary slightly depending on specific cognitive outcomes.^4,6,9,10^ While previous studies have been predominantly conducted in European populations or US populations of European ancestry, several prior studies have demonstrated similar associations in US Hispanic/Latino populations.^13–16^

The majority of previous studies on sleep and cognitive outcomes have utilized self-reported sleep measures. Since polygenic scores (PGS) for sleep are statistically significantly associated with their respective sleep phenotypes^17–20^, they are also useful measures of sleep, especially in studies that have not collected information on sleep via questionnaire. Even in studies with self-reported sleep measures, questions used to ascertain sleep can vary across studies and recalled information may not accurately reflect actual sleep quality or capture all aspects of sleep. In addition, sleep habits are usually not static and can change over the life course.^21^ Therefore, sleep PGS may offer a more and stable marker of sleep. Studying the relationship between sleep PGS and other phenotypes associated with sleep, such as cognitive outcomes, may determine whether the genetics underlying sleep are potentially predictive of related conditions. Furthermore, if sleep PGS is found to be correlated with cognitive functioning, it can then serve as a more stable measure for cognitive impairment risk group identification as compared to self-reported sleep measures.^22^ Especially when studying cognition, individuals who have diminished alertness and concentration due to poor sleep prior to exams often perform badly on cognitive tests temporarily.

Despite the utility of evaluating associations between sleep PGS and cognitive outcomes, few studies have done so. Based on mediation analyses using UK Biobank data (primarily White individuals from UK), Li et al found that a sleep duration PGS was associated with various aspects of cognitive function and brain structure.^12^ Likewise, sleep duration PGS was associated with global cognition and several cognitive domains in the Reference Abilities Neural Network (RANN) and Cognitive Reserve study^23^ and changes in cognition in the Hellenic Longitudinal Investigation of Aging and Diet (HELIAD) study.^24^ Finally, a recent HCHS/SOL study of sleep PGSs and cognitive outcomes reported several associations, including insomnia PGS with lower global cognition and mild cognitive impairment (MCI), sleep duration PGS and excessive daytime sleepiness PGS with MCI.^25^ The present study utilized PGS for sleep traits to assess associations with cognitive function using derived global cognitive outcomes across several multiethnic populations: Atherosclerosis Risk in Communities (ARIC), Bogalusa Heart Study (BHS), Cardiovascular Health Study (CHS), Multi-Ethnic Study of Atherosclerosis (MESA) and Hispanic Community Health Study/Study of Latinos (HCHS/SOL or SOL hereafter). PGSs for sleep traits were constructed based on previously-developed variant and weight files. These PGS cover sleep duration (also short and long sleep), insomnia, excessive daytime sleepiness and OSA, and were applied across cohorts.^17–20^ Finally, we sought to extend previous research by examining associations stratified by sex and race/ethnicity given the differential sleep and cognitive aging patterns across sex and race groups.^26–28^ By comprehensively examining associations between sleep PGS and cognitive outcomes, we sought to characterize the common genetic risk underlying sleep and cognitive function and assess whether relationships between genetic risk for sleep and cognitive function vary across populations. This is the first and largest community-based multi-cohorts study quantifying the associations of PGS for multiple sleep traits with risk of prevalent poor cognitive function, cognitive change and risk of incident poor cognitive function.

## 2. Methods

### 2.1 Study population

We included five US-based longitudinal prospective cohorts that have previously collected participants information on cognitive functioning for at least two visits and with available genotyping data: ARIC, BHS, CHS, MESA, and SOL. The design and details of each cohort are described in **Appendix 1** in supporting information. Here, we used data from two visits, one referred to as baseline (first selected visit) and the other referred as follow-up (subsequent to the first selected visit) . For each cohort, we selected the first available visit with comprehensive cognitive tests as our baseline, and chose its subsequent visit as our follow-up visit (except for CHS, where the follow up visit was selected to enable a follow-up period of at least 5 years). Details on the specific visits treated as baseline and follow-up for each cohort are displayed in **Figure 1**. For each cohort, only participants with complete information on sleep PGS and baseline global cognitive function measure (defined below in Section 2.2) were included.

**Figure 1.**
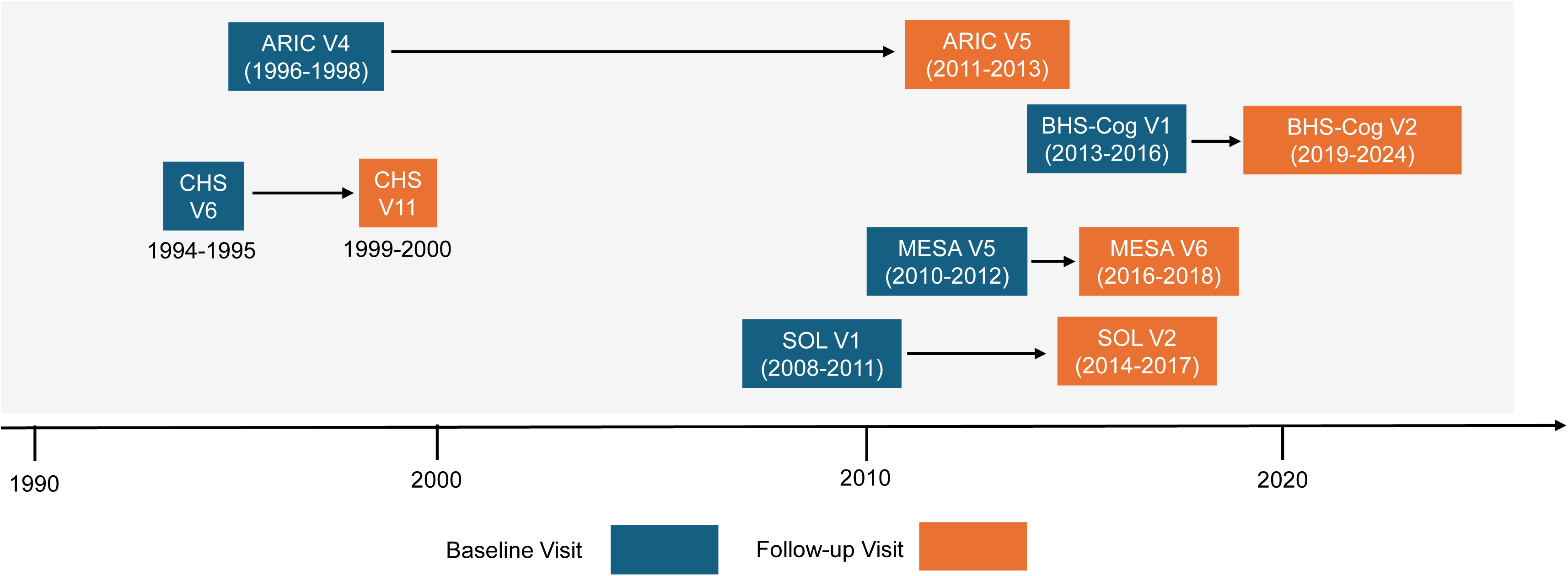
Distribution of Timelines for Selected Study Visits of Included Cohorts. The X axis is the timeline in calendar years. ARIC: Atherosclerosis Risk in Communities study, participants with prevalent stroke at the baseline visit, or prevalent stroke and/or prevalent dementia at the follow-up visit were excluded prior to the generation of the GCF variable; BHS-Cog: Bogalusa Heart Study – Cognition Follow-up ; CHS: Cardiovascular Health Study; MESA: Multi-Ethnic Study of Atherosclerosis; SOL: Hispanic Community Health Study/Study of Latinos.

### 2.2 Assessment of cognitive function

For each cohort, we included at least three cognitive function tests to derive global cognitive function (GCF) measures used for the association analyses with sleep PGSs. Our approach follows similar methodology to that previously established by the Cohorts for Heart and Aging Research in Genomic Epidemiology (CHARGE) consortium, where they used the same baseline GCF measure as the outcome variable in meta-analyses of genome-wide association studies (GWAS).^29,30^ Details on the cognitive function tests in each cohort and their related cognitive domains are listed in **Table S1** in supporting information. First, for each cohort, we derived a baseline GCF measure by applying principal components analysis (PCA)^31,32^ to the standardized (z-scored) individual cognitive functioning tests. Prior to PCA, all cognitive tests were oriented so that higher scores imply better cognitive performance. Baseline GCF was defined as the standardized first principal component (PC1) from the PCA. Higher GCF indicates better cognition. We then derived GCF at follow-up based on the baseline-standardized individual cognitive tests from the follow-up . Here, baseline-standardized means that when standardizing the follow-up individual test scores, and later when standardizing PC1, we used the tests’ (and PC’s) means and SDs computed from the baseline exam. We then used the follow-up and baseline GCF to create a GCF change variable, defined as the GCF value at follow-up minus the GCF value at baseline. We constructed a binary measure for having “poor GCF” or not at baseline in each cohort as whether the individual had a baseline age-adjusted GCF of 1 SD below the distribution median. We additionally constructed a binary outcome variable measuring whether the individual was an incident case of poor GCF at the follow-up visit among participants who were not categorized as having poor GCF at baseline following the same approach for poor GCF at baseline. In summary, we used four global cognitive function measures as our main outcomes: 1) baseline global cognitive function (GCF); 2) poor baseline GCF; 3) GCF change from baseline to follow-up visit; 4) incident poor GCF. The workflow for the derivation of the GCF measures is pictured in **Figure S1** in supporting information.

### 2.3 PGS for sleep traits

We constructed PGSs for the following six sleep traits across the five cohorts: insomnia, long sleep, short sleep, obstructive sleep apnea (OSA), sleep duration and excessive daytime sleepiness. Sleep phenotype definitions in the GWAS studies used as sources for PGS development are documented in **Table S2** in supporting information. This table also reports the GWAS populations, samples sizes, and publications. OSA cases were identified based on clinical diagnoses from leveraged electronic health records. All other sleep traits used in the GWAS studies were self-reported. In ARIC, genotyping was performed using the Affymetrix GeneChip SNP Array 6.0.^33^ Imputation was subsequently carried out using the NHLBI Trans-Omics for Precision Medicine (TOPMed) 2.0 reference panel. Chip-based genotyping was selected here for ARIC due to a larger sample size as compared to whole genome sequencing (WGS). In BHS, genotyping was conducted using Illumina Human610 BeadChip described previously.^34–37^ Genotypes were also subsequently imputed to the TOPMed 2.0 reference panel. For CHS and MESA, WGS genetic data from TOPMed Freeze 10b was obtained. For SOL, we used a combination of WGS from TOPMed (N = 13,025) and imputed data (for a small subset of individuals for which WGS was not available, N = 1,083). For imputed data, genotyping was performed over the Illumina HumanOmni2.5 8 BeadChip and TOPMed 3.0 reference panel was used for imputation. Additional details of genotyping in SOL can be found in previous publication.^38^ Further information on sample quality control, imputation and ancestry genetic PCs generation for each cohort can be found in **Appendix 2** in supporting information.

PGSs for OSA were previously developed by our group, which include two versions (BMI adjusted and unadjusted in GWAS).^19^ We used the PGSs variant weights files generated from this prior study to construct OSA PGSs in each cohort. For other non-OSA traits, we used PRS weights developed using PRS-CS^39^ as detailed in Wyss, et al. 2026,^40^ and based on summary statistics from GWAS of the relevant sleep trait conducted over UK Biobank European British participants.^17,18,20^ Either PLINK or PRSice (without additional clumping or thresholding) was used to construct the PGSs for all sleep traits based on the weights files for each cohort.^41–43^ Before modeling, all PGSs were scaled, separately in each cohort, so that the final effect estimates are calculated as effect sizes per 1 SD increase in each sleep PGS. We estimated Pearson’s correlation coefficients across sleep PGSs in all cohorts.^44^

### 2.4 Statistical analyses

We performed the primary association analyses between each sleep PGS and each global cognitive function measure (GCF at baseline, poor GCF at baseline, GCF change, and incident poor GCF at follow-up) using generalized linear models (GLMs) in each cohort. For SOL, survey-based GLM models incorporating sampling weights were applied to account for the survey sampling design, including stratification, clustering, probability sampling and non- response as detailed in prior publications.^45,46^ For models with GCF and poor GCF at baseline, we adjusted for self-reported age (continuous: in years), self-reported sex (categorical: females, males), self-reported race/ethnicity background (categorical as described in study population section: Hispanic/Latino background for SOL), study locations/centers (categorical as described in study population section), genetic PCs (separate continuous terms for each PC) and self-reported education (categorical: < 12, = 12 and > 12 years) at baseline as primary covariates. For models with GCF change, we additionally adjusted for years between the two visits (continuous: between follow-up visit and baseline) apart from the previous covariates. For models with incident poor GCF at follow-up, we included an offset term of the natural log of the years between the two visits as the follow-up period in a quasi-Poisson regression for estimating incident rate ratio. We meta-analyzed findings across the five cohorts (ARIC, BHS, CHS, MESA, SOL) using inverse-variance fixed-effects meta-analysis^47,48^, and reported the meta-analyzed effect estimate, standard error (SE), p value for Cochran’s Q test, I-squared (I²) (as a measure for effect heterogeneity across the cohort-specific results) and analytical sample size. For all models, we reported differences in baseline GCF or GCF change, odds ratio (OR) of having poor GCF at baseline or incident rate ratio (IRR) of having poor GCF at the follow-up visit, along with their 95% confidence intervals (95% CI) per 1 SD increase in each sleep PGS. We reported statistically significant associations based on a two-sided p<0.05, and marginally significant (suggestive) associations based on a two-sided p<0.10. In addition, we reported point estimates, their 95% CIs along with p values to assist the interpretation of the overall association strength combining magnitude, precision, and statistical evidence.

We conducted a few secondary analyses to assess the robustness of our primary findings and to look into potential reasons for heterogeneity in results across cohorts by 1) first, additionally adjusting for APOE e4 allele status (continuous: having 0, 1, 2 copies of APOE e4 allele); 2) meta-analyzing results from younger cohorts (BHS and SOL, average age at first selected visit <60 yrs.), and from older cohorts (ARIC, MESA and CHS, average age at first selected visit ≥60 yrs.); 3) running stratified analyses by sex to identify potential sex differences; 4) running stratified analyses by race/ethnicity to identify potential differences by race/ethnicity backgrounds. In race/ethnic stratified analysis, we considered White, Black, and Hispanic/Latino groups, but did not study associations in Chinese-only analysis (available from MESA) due to limited sample size. As ad-hoc analyses, heterogeneity Cochran’s Q test was applied to sex-stratified and race-stratified meta-analyzed estimates to further detect statistically significant (Q test p <0.05) association heterogeneity across sex and race/ethnicity groups. All analyses were conducted using R environment version 4.2.2.

## 3. Results

### 3.1 Participants characteristics

Cohort-specific participants characteristics can be found in **Table 1**. The study included 23,781 participants from five contributing observational cohorts: ARIC (N = 8,086), SOL (N = 8,069), BHS (N = 1,078), CHS (N = 2,660) and MESA (N = 3,888). On average, participants ages ranged from 48.2 yrs (BHS) to 75.4 yrs (CHS) at the baseline visit. Percent female ranged from 52.8% in MESA to 61.5% in SOL. ARIC and BHS included White and Black participants, CHS included White, Black participants and participants of other races, SOL included Hispanic/Latino participants, and MESA included White, Black, Hispanic, and Chinese participants. **Figure 1** depicts the selected baseline and follow-up visits and their timelines in each of the cohorts. In addition, we observed moderate to high correlations between PGSs for short sleep and sleep duration, as well as high correlation between BMI-adjusted and -unadjusted OSA PGSs. In BHS, we observed high correlation between PGS for OSA (BMI-adjusted and unadjusted), short sleep and sleep duration. For other traits, low to moderate correlations were observed. Further details can be found in **Table S3** in supporting information.

**Table 1.**
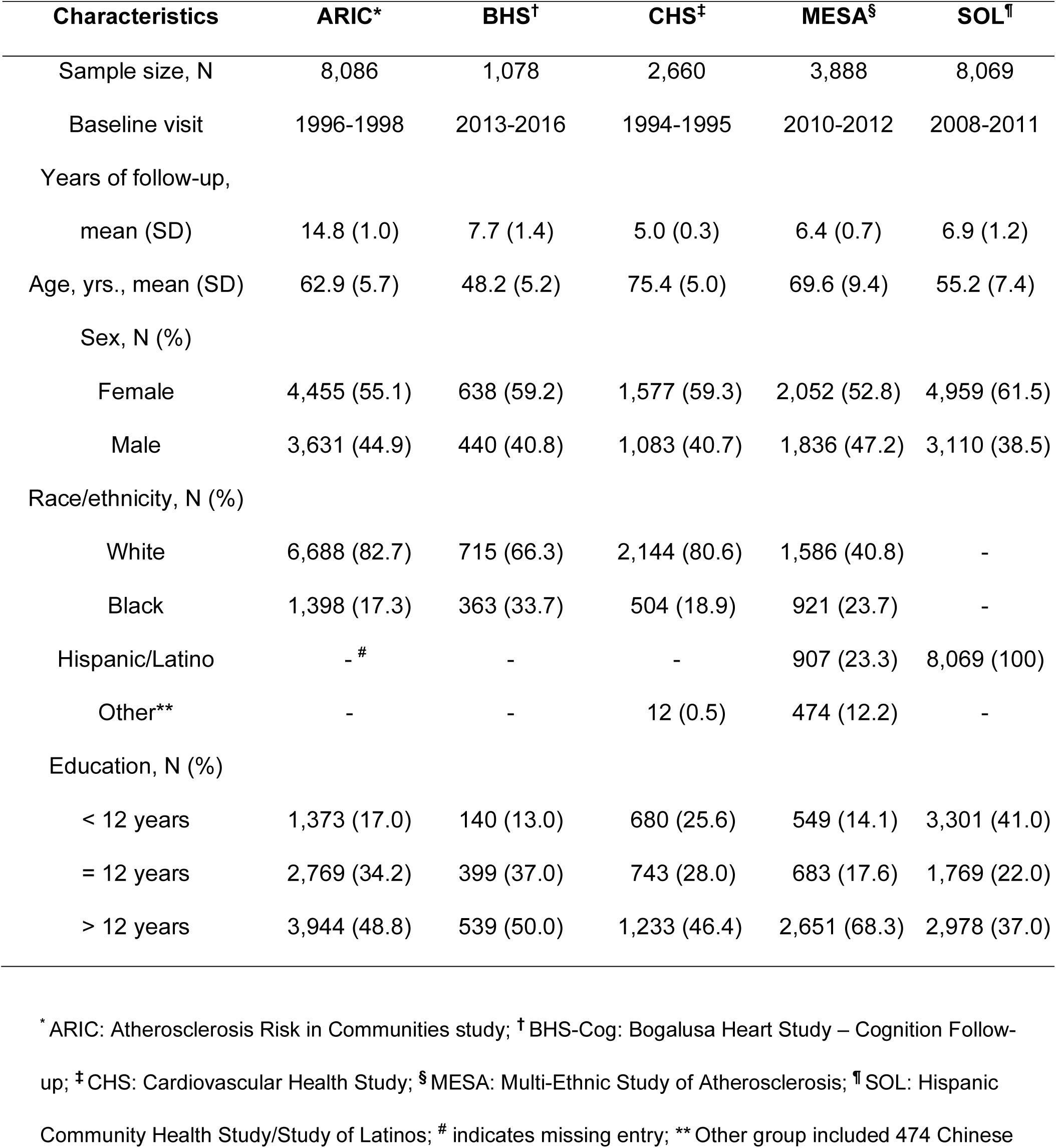
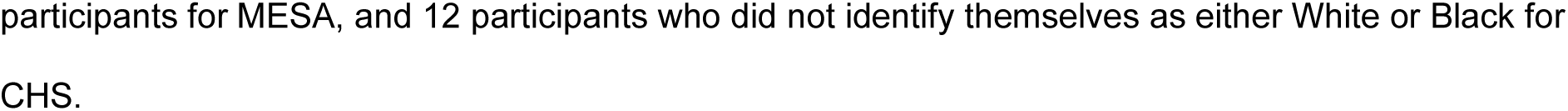
Cohort-specific participants characteristics at first selected visit (treated as baseline) with available polygenic scores (PGS) for sleep traits and derived global cognitive function at baseline.

| **Characteristics** | **ARIC\*** | **BHS<sup>†</sup>** | **CHS<sup>‡</sup>** | **MESA<sup>§</sup>** | **SOL<sup>¶</sup>** |
| --- | --- | --- | --- | --- | --- |
| Sample size, N | 8,086 | 1,078 | 2,660 | 3,888 | 8,069 |
| Baseline visit | 1996-1998 | 2013-2016 | 1994-1995 | 2010-2012 | 2008-2011 |
| Years of follow-up, |  |  |  |  |  |
| mean (SD) | 14.8 (1.0) | 7.7 (1.4) | 5.0 (0.3) | 6.4 (0.7) | 6.9 (1.2) |
| Age, yrs., mean (SD) | 62.9 (5.7) | 48.2 (5.2) | 75.4 (5.0) | 69.6 (9.4) | 55.2 (7.4) |
| Sex, N (%) |  |  |  |  |  |
| Female | 4,455 (55.1) | 638 (59.2) | 1,577 (59.3) | 2,052 (52.8) | 4,959 (61.5) |
| Male | 3,631 (44.9) | 440 (40.8) | 1,083 (40.7) | 1,836 (47.2) | 3,110 (38.5) |
| Race/ethnicity, N (%) |  |  |  |  |  |
| White | 6,688 (82.7) | 715 (66.3) | 2,144 (80.6) | 1,586 (40.8) | - |
| Black | 1,398 (17.3) | 363 (33.7) | 504 (18.9) | 921 (23.7) | - |
| Hispanic/Latino | - <sup>#</sup> | - | - | 907 (23.3) | 8,069 (100) |
| Other** | - | - | 12 (0.5) | 474 (12.2) | - |
| Education, N (%) |  |  |  |  |  |
| < 12 years | 1,373 (17.0) | 140 (13.0) | 680 (25.6) | 549 (14.1) | 3,301 (41.0) |
| = 12 years | 2,769 (34.2) | 399 (37.0) | 743 (28.0) | 683 (17.6) | 1,769 (22.0) |
| > 12 years | 3,944 (48.8) | 539 (50.0) | 1,233 (46.4) | 2,651 (68.3) | 2,978 (37.0) |

\* ARIC: Atherosclerosis Risk in Communities study; <sup>†</sup> BHS-Cog: Bogalusa Heart Study – Cognition Follow-up; <sup>‡</sup> CHS: Cardiovascular Health Study; <sup>§</sup> MESA: Multi-Ethnic Study of Atherosclerosis; <sup>¶</sup> SOL: Hispanic Community Health Study/Study of Latinos; <sup>#</sup> indicates missing entry; \*\* Other group included 474 Chinese
participants for MESA, and 12 participants who did not identify themselves as either White or Black for CHS.

### 3.2 Overall associations of sleep PGSs with cognitive outcomes

We conducted cohort-specific association analyses and meta-analysis on the associations of sleep PGSs with four global cognitive function measures. **Figure 2 and 3** visualize the corresponding cohort-specific and meta-analysis findings for self-reported sleep PGS and OSA PGSs, respectively. Final meta-analysis showed that on average, each 1 SD increase in long sleep PGS was associated with lower baseline global cognitive function (meta-analyzed estimate = -0.02 SD, 95% CI: -0.03 to 0.00, p = 0.01) with no significant heterogeneity across the five cohorts (across-cohorts *P* for Cochran’s Q test = 0.90; I² < 0.01). The long sleep PGS was also associated with higher risk of poor GCF (> 1 SD below the distribution median) at baseline (meta-analyzed OR = 1.04, 95% CI: 1.00 to 1.09, p = 0.06) with no significant heterogeneity across the five cohorts (across-cohorts *P* for Cochran’s Q test = 0.49; I² < 0.01). In addition, BMI-adjusted OSA PGS was linked to higher risk of poor GCF at baseline (meta-analyzed OR = 1.11, 95% CI: 1.00 to 1.22, p = 0.04) per 1 SD increase in PGS with no significant heterogeneity across the cohorts (across-cohorts *P* for Cochran’s Q test = 0.86; I² < 0.01).

**Figure 2.**
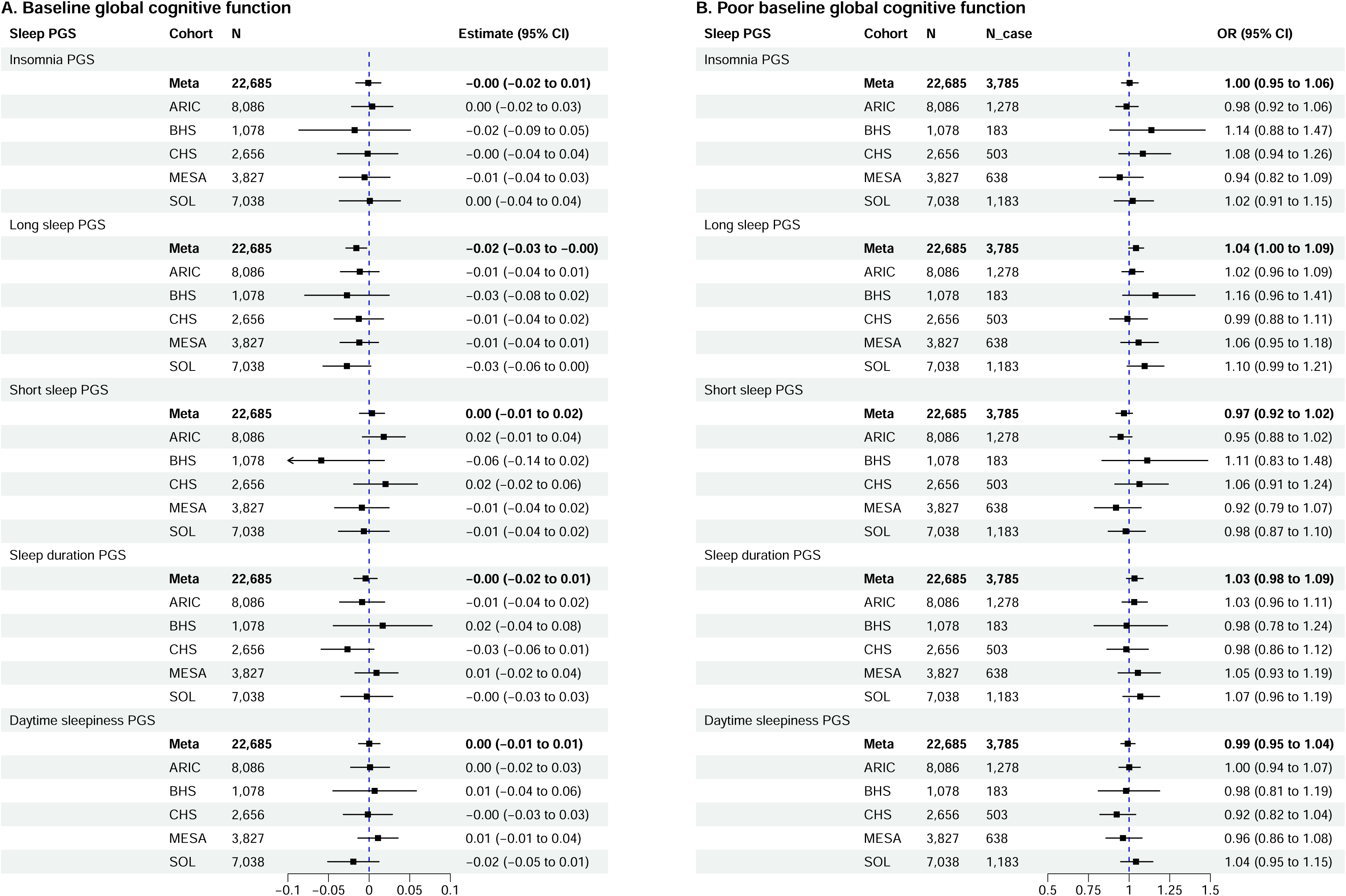
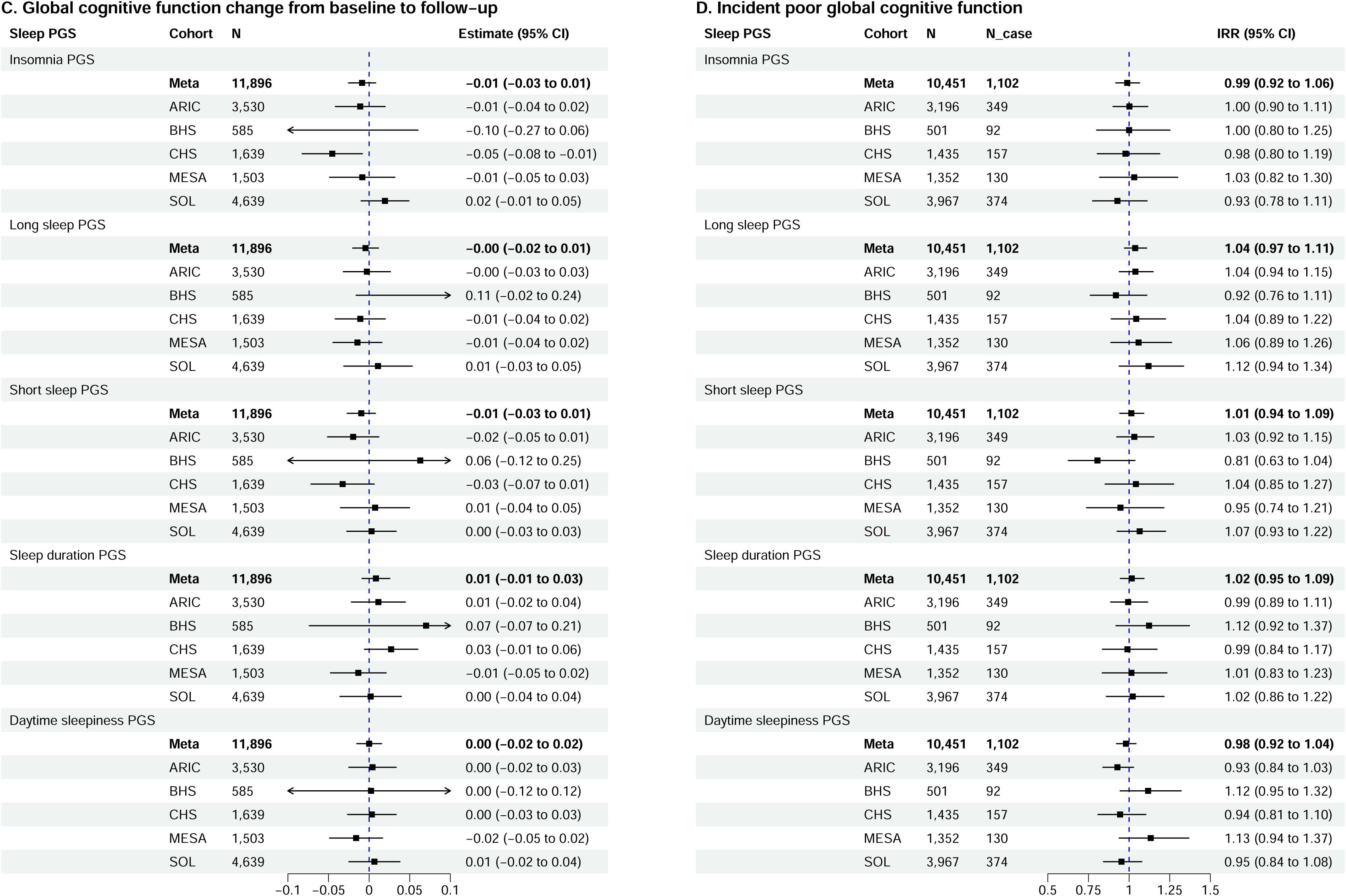
Associations of Polygenic Scores (PGS) for Sleep Traits (non-Obstructive Sleep Apnea) with Global Cognitive Outcomes in Each Cohort and from Meta-Analysis. Covariates adjusted include age, sex, race/ethnicity background, study locations/centers, genetic principal components (PCs) and education at baseline. For models with global cognitive function (GCF) change from baseline to follow-up, years between baseline and follow-up visits was adjusted apart from the previous covariates. For models with incident risk at follow-up, an offset term of the natural log of the years between baseline and follow-up visits was included. N: sample size included; N_case: number of baseline poor GCF or incident poor GCF cases; Estimate: point difference in GCF or GCF change per 1 SD increase in a PGS; 95% CI: 95% confidence interval; OR: odds ratio; IRR: incident rate ratio; Meta: meta-analysis; ARIC: Atherosclerosis Risk in Communities study; BHS-Cog: Bogalusa Heart Study – Cognition Follow-up ; CHS: Cardiovascular Health Study; MESA: Multi-Ethnic Study of Atherosclerosis; SOL: Hispanic Community Health Study/Study of Latinos.

**Figure 3.**
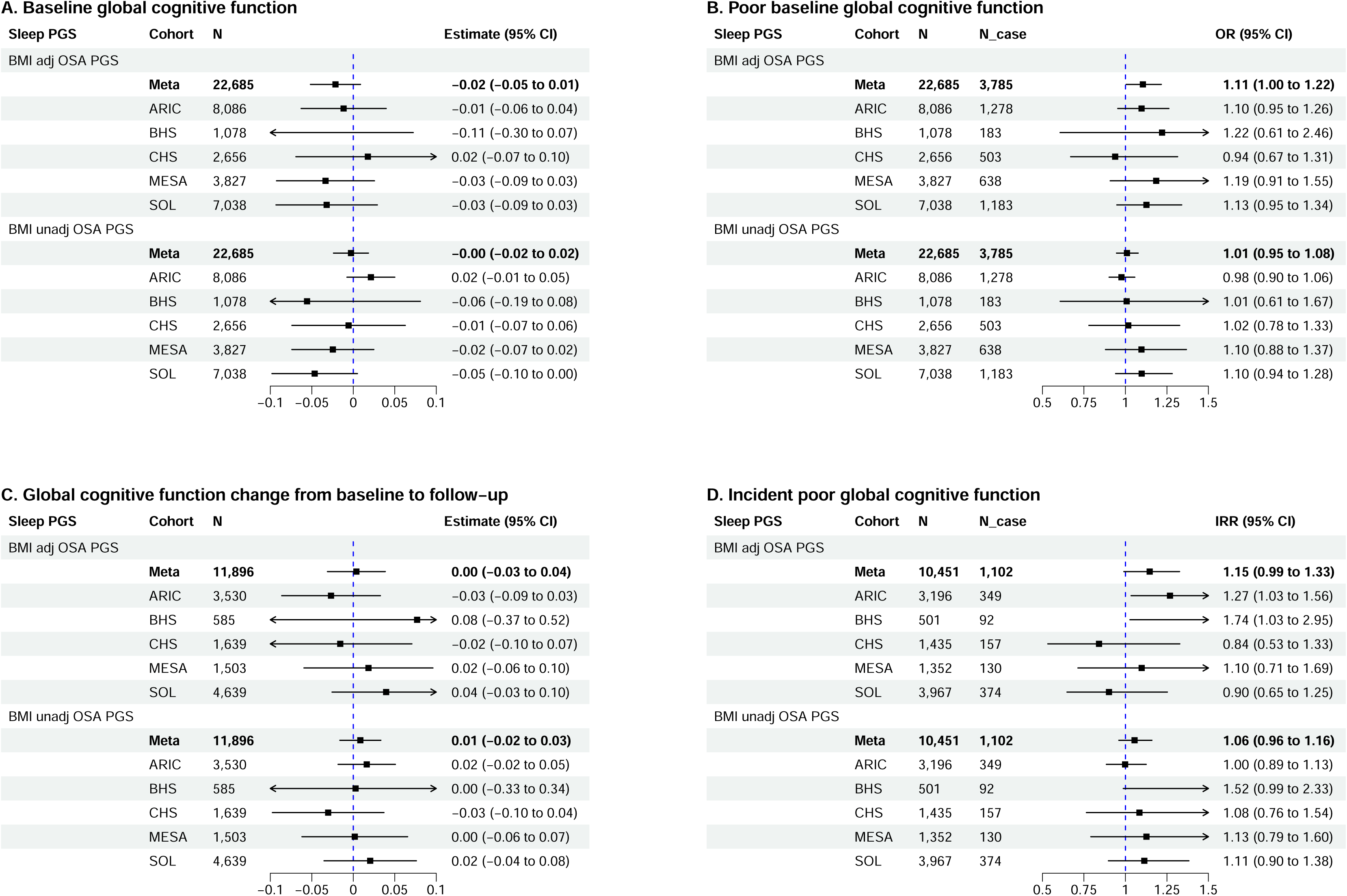
Associations of Polygenic Scores (PGS) for Obstructive Sleep Apnea (OSA) with Global Cognitive Outcomes in Each Cohort and from Meta-Analysis. Covariates adjusted include age, sex, race/ethnicity background, study locations/centers, genetic principal components (PCs) and education at baseline. For models with global cognitive function (GCF) change from baseline to follow-up, years between baseline and follow-up visits was adjusted apart from the previous covariates. For models with incident risk at follow-up, an offset term of the natural log of the years between baseline and follow-up visits was included. N: sample size included; N_case: number of baseline poor GCF or incident poor GCF cases; Estimate: point difference in GCF or GCF change per 1 SD increase in a PGS; 95% CI: 95% confidence interval; OR: odds ratio; IRR: incident rate ratio; Meta: meta-analysis; ARIC: Atherosclerosis Risk in Communities study; BHS-Cog: Bogalusa Heart Study – Cognition Follow-up ; CHS: Cardiovascular Health Study; MESA: Multi-Ethnic Study of Atherosclerosis; SOL: Hispanic Community Health Study/Study of Latinos; BMI adj: BMI adjusted; BMI unadj: BMI unadjusted.

Overall, there were no statistically significant associations between any sleep PGS with GCF change from baseline to follow-up based on the meta-analysis. This was also largely true across the five cohorts except for insomnia PGS’s association with reduced GCF from baseline to follow-up in CHS (point estimate = -0.05 SD, 95% CI: -0.08 to - 0.01, p = 0.02). In addition, we observed a marginal association between OSA PGS and incident poor GCF risk at follow-up based on meta-analysis. Each 1 SD increase in BMI-adjusted OSA PGS was associated with increased risk of incident poor GCF at follow-up (meta-analyzed IRR = 1.15, 95% CI: 0.99 to 1.33, p = 0.07) with moderate heterogeneity across the cohort-specific findings (across-cohorts *P* for Cochran’s Q test = 0.12; I² = 44.57). Related meta-analysis effect estimates, 95% CI, *P* for Q test, I² and total sample size are presented in **Table S4** in supporting information. We did not observe significant associations of any cognitive outcomes with PGSs for insomnia, short sleep, sleep duration, sleepiness or BMI unadjusted OSA in the meta-analysis of total population. Further accounting for APOE e4 carrier status did not alter these findings **(Table S5).**

Secondary analyses meta-analyzing younger (BHS and SOL) vs older cohorts (ARIC, MESA and CHS) suggests that the inverse associations between long sleep PGS and baseline cognition are more profound in younger cohorts. In addition, each 1 SD increase in insomnia PGS was associated with a GCF change of -0.02 SD (95% CI: - 0.04 to 0.00, p = 0.05) from baseline to follow-up in the older cohorts. Further details can be viewed in **Table S6.** More information regarding the total non-stratified findings in each cohort can be found in **Table S7**.

### 3.3 Sex-specific findings

**Figure 4** displays sex-specific effect estimates from meta-analysis (A.GCF at Baseline; B. Poor GCF at Baseline; C. GCF Change; D. Incident Poor GCF at Follow-up). The results suggest that, consistent with the main findings, higher genetic risk for long sleep was associated with lower baseline GCF among both males and females with no effect heterogeneity (across-groups *P* for Cochran’s Q test = 0.83, I² < 0.01). Specifically, on average, each 1 SD increase in long sleep PGS was associated with lower baseline GCF in females (meta-analyzed estimate = -0.01 SD, 95% CI: -0.03 to 0.00, p = 0.11), and lower baseline GCF in males with similar effect size (meta-analyzed estimate = -0.02 SD, 95% CI: -0.04 to 0.00, p = 0.07). No associations were observed with GCF change from baseline to follow-up among either group. In addition, among females, each 1 SD increase in OSA PGS (after accounting for BMI) was associated with a 22% increased risk of developing incident poor GCF at the follow-up visit (meta-analyzed IRR = 1.22, 95% CI: 1.02 to 1.45, p = 0.03) with strong heterogeneity observed across the cohorts (within female *P* for Cochran’s Q test < 0.01; I² = 73.79). We did not observe such association among males (meta-analyzed IRR = 1.11, 95% CI: 0.91 to 1.36, p = 0.31) though similar effect heterogeneity was observed among males across cohorts (within male *P* for Cochran’s Q test = 0.03; I² = 62.99). In addition, no statistically significant effect heterogeneity was observed when comparing females and males for the associations between BMI adjusted OSA PGS and incident poor GCF (across-groups *P* for Cochran’s Q test = 0.50, I² < 0.01). More information regarding the sex-specific findings from meta-analysis and in each cohort can be found in **Table S8** and **Table S9**.

**Figure 4.**
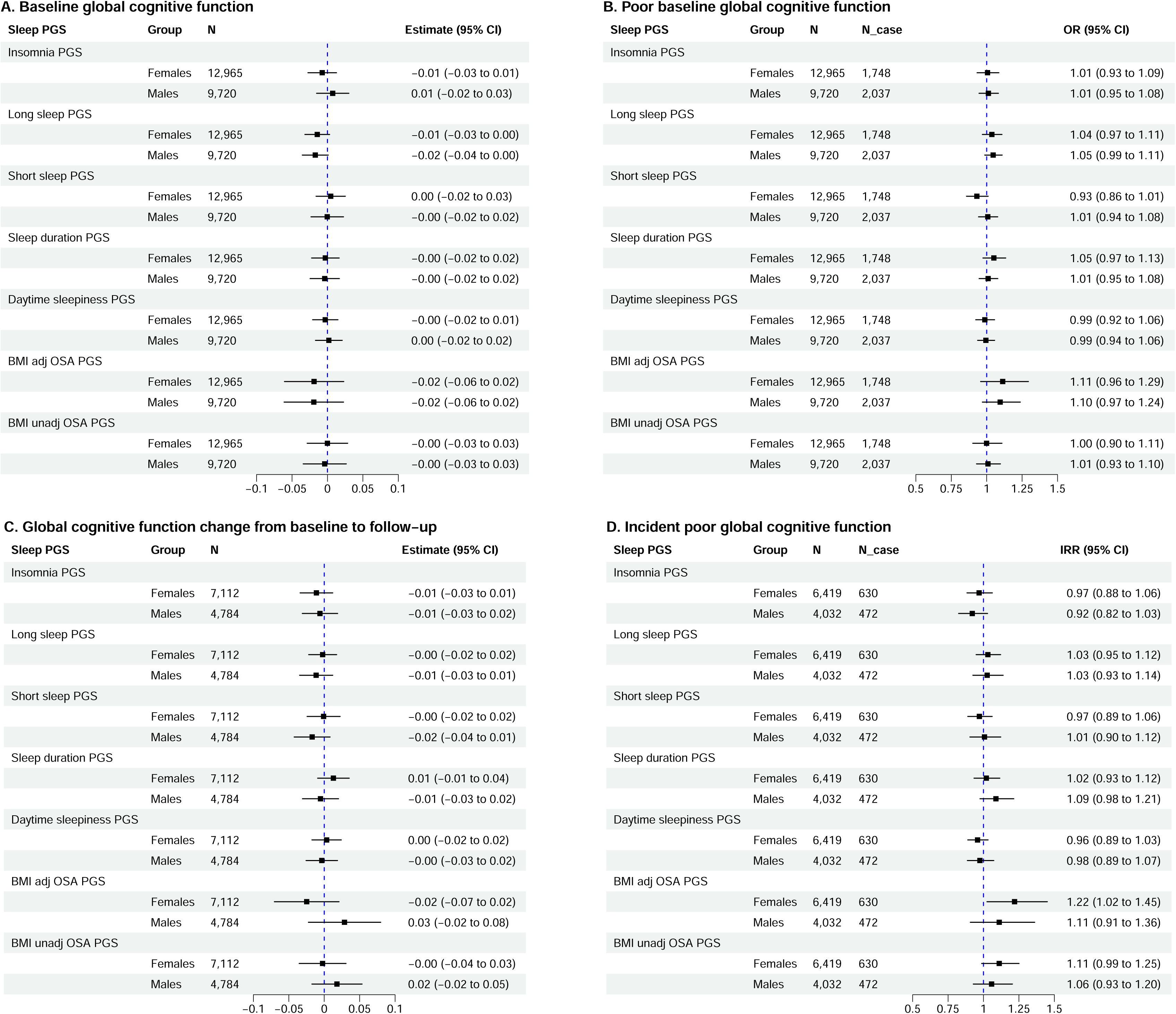
Sex-specific Associations of Polygenic Scores (PGS) for Sleep Traits with Global Cognitive Outcomes in Each Cohort and from Meta-Analysis. Covariates adjusted include age, race/ethnicity background, study locations/centers, genetic principal components (PCs) and education at baseline. For models with global cognitive function (GCF) change from baseline to follow-up, years between baseline and follow-up visits was adjusted apart from the previous covariates. For models with incident risk at follow-up, an offset term of the natural log of the years between baseline and follow-up visits was included. N: sample size included; N_case: number of baseline poor GCF or incident poor GCF cases; Estimate: point difference in GCF or GCF change per 1 SD increase in a PGS; 95% CI: 95% confidence interval; OR: odds ratio; IRR: incident rate ratio; Meta: meta-analysis; ARIC: Atherosclerosis Risk in Communities study; BHS-Cog: Bogalusa Heart Study – Cognition Follow-up; CHS: Cardiovascular Health Study; MESA: Multi-Ethnic Study of Atherosclerosis; SOL: Hispanic Community Health Study/Study of Latinos; BMI adj: BMI adjusted; BMI unadj: BMI unadjusted.

### 3.4 Race/ethnicity-specific findings

**Figure 5** shows effect estimates from race-specific meta-analysis (A.GCF at Baseline; B. Poor GCF at Baseline; C. GCF Change; D. Incident Poor GCF at Follow-up). The overall associations we observed between long sleep PGS and lower baseline global cognitive function were still present among White and Hispanic individuals. On average, each 1 SD increase in PGS for long sleep was associated with -0.02 SD lower baseline GCF (95% CI: -0.04 to 0.00, p = 0.03) among White individuals and -0.03 SD lower baseline GCF (95% CI: -0.05 to 0.00, p = 0.05) among Hispanic individuals. Long sleep PGS was also marginally associated with higher risk of poor GCF at baseline for both White (OR: 1.04, 95% CI: 0.98 to 1.11, p = 0.17) and Hispanic (OR: 1.08, 95% CI: 0.99 to 1.18, p = 0.08) individuals, as well as increased risk of incident poor GCF at follow-up for White individuals (IRR: 1.08, 95% CI: 0.99 to 1.17, p = 0.08). However, there were null associations for long sleep PGS on cognitive outcomes among Black individuals. We observed evidence of statistically significant effect heterogeneity when comparing across White, Black and Hispanic groups between long sleep PGS and incident poor GCF (across-groups *P* for Cochran’s Q test < 0.05, I² = 66.74). In addition, each 1 SD increase in BMI-adjusted OSA PGS was associated with a 12% increased risk of developing incident poor GCF at follow-up (meta-analyzed IRR = 1.12, 95% CI: 1.01 to 1.25, p = 0.03) among White individuals, and 20% higher risk of being poor GCF at baseline (meta-analyzed OR = 1.20, 95% CI: 1.02 to 1.40, p = 0.03) among Hispanic individuals. However, the observed heterogeneity by race/ethnicity background for the associations between BMI-adjusted OSA PGS and incident poor GCF was moderate but not statistically significant (across-groups *P* for Cochran’s Q test = 0.16, I² = 45.59). Stratification by race/ethnicity background also revealed a few potential race-specific associations. For example, among White individuals, higher genetic risk for insomnia was associated with global cognitive function decline from baseline to follow-up (meta-analyzed estimate = -0.02 SD, 95% CI: -0.04 to 0.00, p = 0.05, per 1 SD increase in insomnia PGS). Among Black individuals, higher genetic risk for short sleep was associated with global cognitive function decline from baseline to follow-up (meta-analyzed estimate = -0.06 SD, 95% CI: -0.11 to -0.01, p = 0.03, per 1 SD increase in short sleep PGS). However, heterogeneity Q test comparing the above race-specific findings did not reach statistical significance for detecting a true heterogeneity (insomnia-GCF change across-groups *P* for Cochran’s Q test = 0.06, I² = 63.65; short sleep – GCF change across-groups *P* for Cochran’s Q test = 0.13, I² = 50.45). More information regarding the race/ethnic-specific findings from meta-analysis and in each cohort can be found in **Table S10** and **Table S11**.

**Figure 5.**
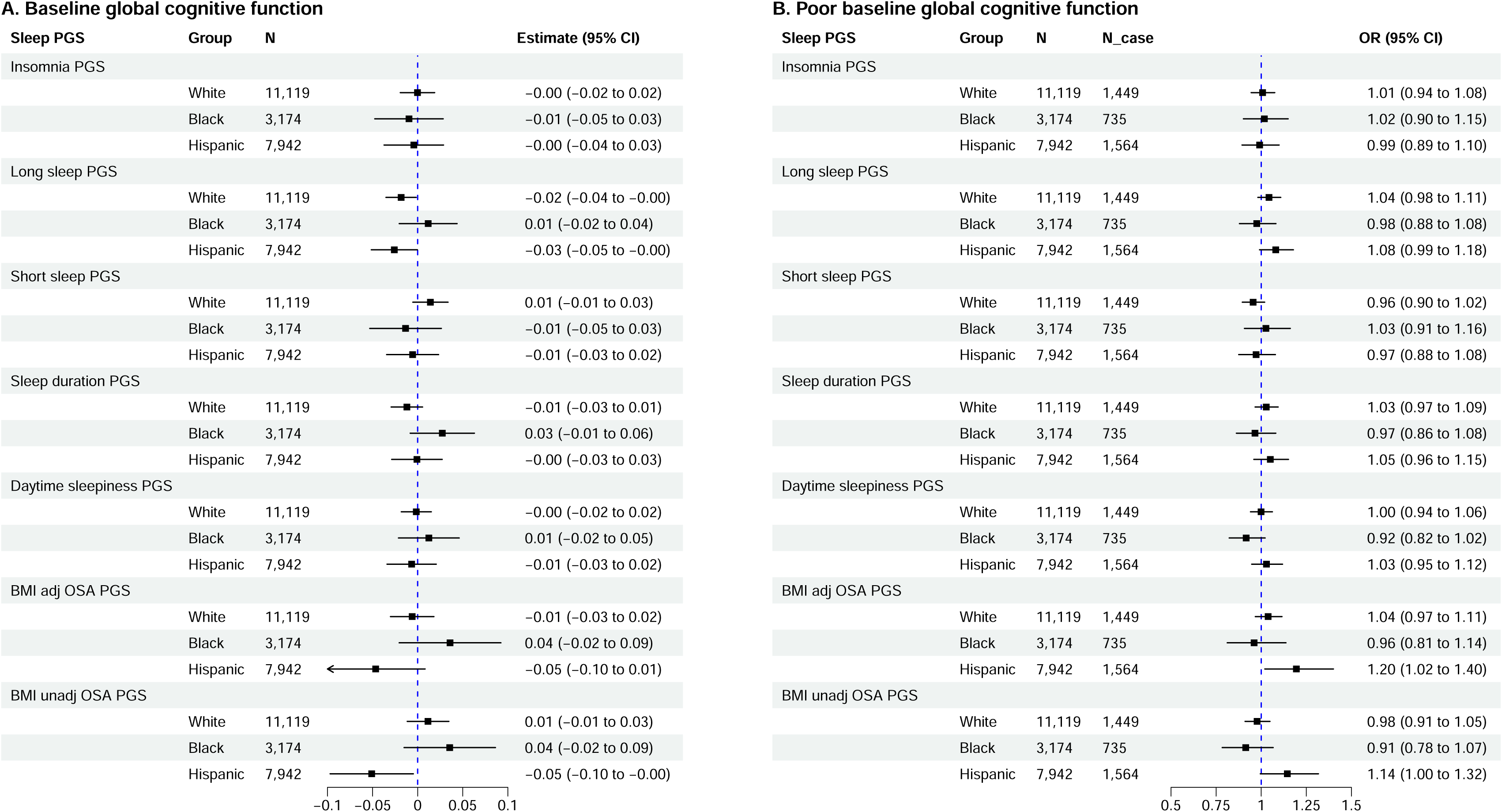
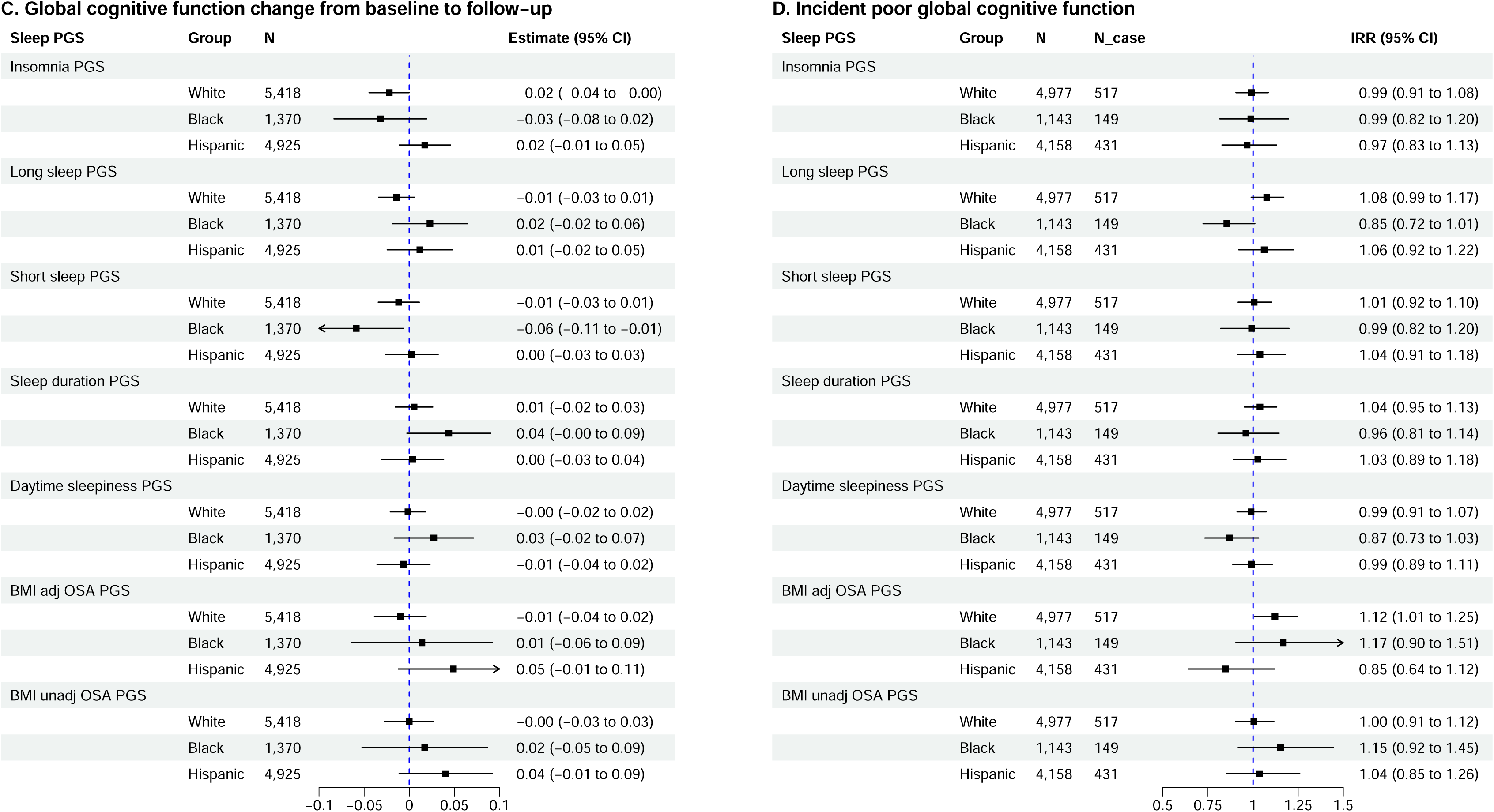
Race/ethnicity-specific Associations of Polygenic Scores (PGS) for Sleep Traits with Global Cognitive Outcomes in Each Cohort and from Meta-Analysis. Covariates adjusted include age, sex, study locations/centers, genetic principal components (PCs) and education at baseline. For models with global cognitive function (GCF) change from baseline to follow-up, years between baseline and follow-up visits was adjusted apart from the previous covariates. For models with incident risk at follow-up, an offset term of the natural log of the years between baseline and follow-up visits was included. N: sample size included; N_case: number of baseline poor GCF or incident poor GCF cases; Estimate: point difference in GCF or GCF change per 1 SD increase in a PGS; 95% CI: 95% confidence interval; OR: odds ratio; IRR: incident rate ratio; Meta: meta-analysis; ARIC: Atherosclerosis Risk in Communities study; BHS-Cog: Bogalusa Heart Study – Cognition Follow-up ; CHS: Cardiovascular Health Study; MESA: Multi-Ethnic Study of Atherosclerosis; SOL: Hispanic Community Health Study/Study of Latinos; BMI adj: BMI adjusted; BMI unadj: BMI unadjusted.

## 4. Discussion

Meta-analyzing evidence across five population-based observational cohorts, we found robust evidence supporting that higher polygenic risk for long sleep and BMI-adjusted OSA was associated with lower level of global cognitive function. Further stratification by sex showed that the association between long sleep PGS and baseline GCF was present among both males and females, and there was no statistically significant evidence of modification by sex. We also observed some differences by race/ethnicity background on the association strength, including in associations of long sleep PGS with baseline GCF (non-significant in Black individuals), of BMI-adjusted OSA PGS with baseline GCF (stronger association for Hispanic individuals), of insomnia PGS with GCF change (significant only in White individuals) and of short sleep PGS with GCF change (significant only in Black individuals). However, further heterogeneity test did not reveal statistically significant association heterogeneity for the above findings across different race/ethnicity backgrounds. Furthermore, interpretation of the race-specific findings should also take into consideration the substantial smaller sample size of included Black individuals and resulting limited power of analyses for this group, as well as the PGS of self-reported sleep traits being derived from a population of European genetic ancestry. Overall, the developed BMI-adjusted OSA PGS (capturing genetic architecture of OSA conditional on BMI) had stronger associations with cognitive outcomes compared to BMI-unadjusted OSA PGS.

The use of sleep PGS in the study of cognitive functioning among clinical and observational cohort studies is still limited. Our study presents suggestive evidence supporting the association of higher PGSs for long sleep and OSA with worse cognitive outcomes. These associations are consistent with current evidence of non-optimal sleep duration and OSA’s associations with worse cognitive outcomes.^49–55^ Research investigating the associations between sleep PGSs and cognitive outcomes remains limited and applied different approaches for measuring cognition and constructing PGSs, limiting direct evidence comparison. In one study examining the association between sleep duration PGS and cognitive changes over a three-years follow up period in the Greek population, researchers found that higher level of sleep duration PGS (constructed using clumping and thresholding methods via PRSice, based on the same GWAS summary statistics^17^ we used here) was associated with cognitive decline over time, particularly with decreased visual-spatial ability trajectories.^24^ In the UK Biobank, Li, et al found that sleep duration PGS (similarly constructed using clumping and thresholding of the same GWAS summary statistics) was associated with several domains of cognitive function and brain structure.^12^ Despite using the same GWAS study, in this meta-analysis, we did not find an association for sleep duration PGS in the total study sample, perhaps due to different PGS methodology, differences in study populations, and variation in the cognitive measures (individual test scores vs. derived global measures). In another study investigating OSA and neurocognitive characteristics leveraging diverse cohorts in a two-sample bidirectional Mendelian Randomization approach, authors found statistically significant associations between OSA and higher hippocampus volume (potentially related to edema and inflammation) using genome-wide significant SNPs (obtained from Strausz, et al. 2021 GWAS^56^) as instrumental variables for OSA.^57^ The same study further detected bidirectional links, where brain cortical structure, brain subcortical, and cognitive performance are associated with OSA risk. Sleep is important for healthy brain development earlier in life, so that the same sleep related genes that are associated with cognition might also influence cognitive development and functioning later in life.^58^ These findings linking sleep with cognitive health using genetic analysis suggest that there might be common genes playing a role in sleep regulation, brain structure and functioning.^12^

In a prior study led by our group looking at sleep PGSs and measures of cognitive functioning in SOL, PGSs of insomnia, daytime sleepiness and sleep duration were associated with in higher risk of prevalent mild cognitive impairment in the Hispanic/Latino population.^25^ In the current study, within SOL, we only found associations between PGSs of long sleep and OSA, and poorer global cognitive functioning. Possible reasons explaining the discrepancy may include the following. First, here we used derived PCs as the GCF phenotypes to match work performed in the Cohorts for Heart and Aging Research in Genomic Epidemiology (CHARGE) consortium^29,30^, while the earlier study looked at prevalent MCI and averaged cognitive test z scores. Second, we conducted additional investigation of long sleep and short sleep traits in our study while the prior study did not. In addition, there were differences in the construction of PGSs: here, we utilized weights from a recent publication^19^ for OSA PGS based on a mixed-population of MVP and FinnGen while the earlier study referred to the Strausz, et al. 2021 GWAS (based on only FinnGen) ^56^; and previous PGS for self-reported sleep phenotypes relied on the same GWAS, but using the less-powerful clumping-and thresholding approach. Notably, within SOL, the PGSs used in the current study had stronger associations with their phenotypes compared to the older PGSs. Together, we think that the difference in cognitive outcomes is what drives the difference in associations between the two SOL analyses. Overall, we found no significant modification by sex for the observed associations of long sleep and OSA PGSs with global cognitive function at baseline, and a potential moderate modification by race/ethnicity. Additional discussion on the potential association heterogeneity by sex, race/ethnicity group and by older vs. younger cohorts is provided in **Appendix 3** in supporting information.

The findings of this study are strengthened by several study design and analytical strengths. First, the evidence presented in this study covers multiple cohorts with over 20,000 middle-aged to older adults of White, Black, Hispanic or other race/ethnicity background, enhancing power and generalizability. We meta-analyzed evidence from five large observational cohorts with mixed ancestry backgrounds, applying a uniformed measure for global cognitive functioning based on standardized primary PC of test z scores within each cohort, and looking at both BMI-adjusted and BMI-unadjusted OSA PGSs developed using a reference population with mixed race/ethnicity backgrounds. Second, it investigated potential differences by sex and race/ethnicity backgrounds, which can help establish targeted clinical screening and prevention efforts for different demographic groups. Third, the application of polygenic scores to estimate the genetic predisposition for related sleep trait is subject to less confounding from external environmental factors (such as time-varying and unknown personal socioeconomic and behaviors) compared to self-reported sleep quality since the underlying genetic determinants are fixed at birth and rarely change over time.

Our study also has a few limitations. First, we cannot draw firm causal conclusions from the analyses due to the nature of observational cohort study. However, since we study the genetic propensity of a sleep trait which is fixed at one’s birth, the study question itself leads to clear distinction between the time window of exposure and outcome, and therefore increasing our confidence in the causality of the presented evidence. Second, the GWAS summary statistics used for the construction of PGSs for sleep traits (except for OSA) are based on European populations which may not align well with the actual effect sizes among population of other genetic ancestries. Third, there could be selection bias within each cohort after excluding people without genetic data for the construction of sleep PGSs and without cognitive measures. However, since the selection was based on genetic data availability which may not necessarily correlate with levels of sleep PGSs, the influence of the relevant selection bias remains limited. Next, the derived global cognitive outcomes we utilized in this study are based on first principal component (explained over 50% of the total variance in test scores) of the standardized cognitive functioning test scores, which may have limited real-world clinical implications. The cognitive tests also varied slightly among different studies while capturing similar domains. In addition, we referred to the nominal significance level at p<0.05 for the identification of ‘suggestive’ significant meta-analysis findings in this study. No associations (main, by sex and by race/ethnicity) remained significant after applying FDR correction to address potential multiple testing in our analyses. However, multiple testing correction may be overly conservative to identify possible associations of interest for future research in that we are not testing against a single hypothesis of biomarker-outcome association like in large omics association analyses but instead making a few comparisons across different sleep PGSs and cognitive measures, and drawing exploratory conclusions for each pair of comparison.^59^ Further, our hypotheses were grounded in previous literature demonstrating poor sleep association with reduced cognitive function and accelerated cognitive aging.^60–62^ We reported point estimates, their 95% CIs along with p values to assist the interpretation of the overall association strength combining magnitude, precision, and statistical evidence.

In conclusion, leveraging information from five large observational cohorts of middle-aged to older adults, we comprehensively estimated the associations between genetic predisposition risk for multiple sleep traits and global cognitive functioning. We found suggestive evidence supporting that higher genetic risk (higher PGS) for long sleep and OSA are associated with lower global cognitive function. These findings add to the scientific basis for using sleep PGS to study the connections between sleep and cognition among middle-aged and older adults.

## Data Availability

Genetic and phenotype data for the involved cohorts are available through application to the Data Base of Genotypes and Phenotypes (dbGaP; https://dbgap.ncbi.nlm.nih.gov). ARIC data are available in accessions phs000280 and phs000090. BHS data are available in accession phs004173. CHS data are available in accession phs001368 (TOPMed WGS). MESA data are available in accession phs001416 (TOPMed WGS). HCHS/SOL data are available in accessions phs000810 and phs000880.

## Acknowledgements

This work was supported by National Heart, Lung, and Blood Institute (NHLBI) grant R01HL161012 to TS and National Institute on Aging (NIA) grant R01AG80598 to TS.

The Atherosclerosis Risk in Communities (ARIC) study has been funded in whole or in part with Federal funds from the NHLBI, National Institutes of Health, Department of Health and Human Services, under Contract nos. (75N92022D00001, 75N92022D00002, 75N92022D00003, 75N92022D00004, 75N92022D00005). The ARIC Neurocognitive Study is supported by U01HL096812, U01HL096814, U01HL096899, U01HL096902, and U01HL096917 from the NIH (NHLBI, NINDS, NIA and NIDCD). The authors thank the staff and participants of the ARIC study for their important contributions. Funding was also supported by R01HL087641 and R01HL086694; National Human Genome Research Institute contract U01HG004402; and National Institutes of Health contract HHSN268200625226C. Infrastructure was partly supported by Grant Number UL1RR025005, a component of the National Institutes of Health and NIH Roadmap for Medical Research.

The Bogalusa Heart Study (BHS) is primarily supported by long-term funding from the NHLBI. The authors thank the staff and participants of the BHS study for their important contributions. Funding was also supported by R01AG077000 from NIA to TK, and by AHA 25CDA1447844 from American Heart Association to XL.

Whole genome sequencing (WGS) for the Trans-Omics in Precision Medicine (TOPMed) program was supported by the NHLBI. Centralized read mapping and genotype calling, along with variant quality metrics and filtering were provided by the TOPMed Informatics Research Center (3R01HL-117626-02S1). Phenotype harmonization, data management, sample-identity QC, and general study coordination, were provided by the TOPMed Data Coordinating Center (3R01HL-120393-02S1). The authors gratefully acknowledge the studies and participants who provided biological samples and data for TOPMed.

WGS for “NHLBI TOPMed: Multi-Ethnic Study of Atherosclerosis (MESA)” (phs001416.v1.p1) was performed at the Broad Institute of MIT and Harvard (3U54HG003067-13S1). The MESA projects are conducted and supported by NHLBI in collaboration with MESA investigators. Support for MESA is provided by contracts 75N92025D00022, 75N92020D00001, HHSN268201500003I, N01-HC-95159, 75N92025D00026, 75N92020D00005, N01-HC-95160, 75N92020D00002, N01-HC-95161, 75N92025D00024, 75N92020D00003, N01-HC-95162, 75N92025D00027, 75N92020D00006, N01-HC-95163, 75N92025D00025, 75N92020D00004, N01-HC-95164, 75N92025D00028, 75N92020D00007, N01-HC-95165, N01-HC-95166, N01-HC-95167, N01-HC-95168, N01-HC-95169, UL1-TR-000040, UL1-TR-001079, UL1-TR-001420, UL1TR001881, R01AG058969 and R01HL105756. The MESA data collection was also supported by R01HL127659. The authors thank the MESA participants and the MESA investigators and staff for their valuable contributions.

Genome Sequencing for “NHLBI TOPMed: Cardiovascular Health Study (CHS)” (phs001368.v1.p1) was performed at Baylor College of Medicine Human Genome Sequencing Center (3U54HG003273-12S2, HHSN268201500015C, and HHSN268201600033I) and the Broad Institute Genomics Platform (HHSN268201600034I). This CHS research was supported by contracts HHSN268201200036C, HHSN268200800007C, HHSN268201800001C, N01HC55222, N01HC85079, N01HC85080, N01HC85081, N01HC85082, N01HC85083, N01HC85086, 75N92021D00006, and U01HL080295, U01HL130114, R01HL105756 and R01HL172803 from the National Heart, Lung, and Blood Institute (NHLBI), with additional contribution from the National Institute of Neurological Disorders and Stroke (NINDS). Additional support was provided by R01AG023629 from the National Institute on Aging (NIA). A full list of principal CHS investigators and institutions can be found at CHS-NHLBI.org.

The Hispanic Community Health Study/Study of Latinos (HCHS/SOL) is a collaborative study supported by contracts from the NHLBI to the University of North Carolina (75N92025D00001, HHSN268201300001I / N01-HC-65233), University of Miami (75N92025D00009, HHSN268201300004I / N01-HC-65234), Albert Einstein College of Medicine (75N92025D00007, HHSN268201300002I / N01-HC-65235), University of Illinois at Chicago (75N92025D00008, HHSN268201300003I / N01-HC-65236 Northwestern Univ), and San Diego State University (75N92025D00010, HHSN268201300005I / N01-HC-65237). The following Institutes/Centers/Offices have contributed to the HCHS/SOL through a transfer of funds to the NHLBI: National Institute on Minority Health and Health Disparities, National Institute on Deafness and Other Communication Disorders, National Institute of Dental and Craniofacial Research, National Institute of Diabetes and Digestive and Kidney Diseases, National Institute of Neurological Disorders and Stroke, NIH Institution-Office of Dietary Supplements. The HCHS/SOL Genetic Analysis Center at the University of Washington was supported by NHLBI and NIDCR contracts (HHSN268201300005C AM03 and MOD03). WGS data for the TOPMed program was supported by the NHLBI. The Centers for Common Disease Genomics (CCDG) program was supported by NHGRI and NHLBI. Genome sequencing for “NHLBI TOPMed: Hispanic Community Health Study/Study of Latinos” (phs001395.v1.p1) was performed at the Baylor College of Medicine Human Genome Sequencing Center (HHSN268201600033I). The authors thank the staff and participants of the HCHS/SOL study for their important contributions. Funding was also supported by R01AG085320 from NIA to RK.

This manuscript is the result of funding in whole or in part by the National Institutes of Health (NIH). It is subject to the NIH Public Access Policy. Through acceptance of this federal funding, the NIH has been given a right to make this manuscript publicly available in PubMed Central upon the Official Date of Publication, as defined by the NIH. The content is solely the responsibility of the authors and does not necessarily represent the official views of the NIH.

## Consent Statement

All participants gave written informed consent. Study protocols were approved by the institutional review boards (IRB) at participating institutions and study centers.

