## Supplemental materials for "Associations of polygenic scores for sleep traits with cognitive function"

**Supplementary Materials**

**Table of Contents**

**Appendix 1.** Cohort details

**Appendix 2.** Genetic data

**Appendix 3.** Additional discussion

**Table S1.** Cognitive tests used to derive global cognitive outcomes in each cohort

**Table S2**. GWAS studies used for the development of polygenic scores (PGS) for sleep traits

**Table S3.** Pearson correlations of polygenic scores for sleep traits in each cohort

**Table S4.** Meta-analyzed effect estimates and heterogeneity test statistics for the associations of polygenic scores (PGS) for sleep traits and global cognitive outcomes in overall population

**Table S5.** Meta-analyzed effect estimates and heterogeneity test statistics for the associations of polygenic scores (PGS) for sleep Traits and global cognitive outcomes in overall population additionally adjusting for APOE e4 allele status

**Table S6.** Meta-analyzed effect estimates and heterogeneity test statistics for the associations of polygenic scores (PGS) for sleep traits and global cognitive outcomes in overall population comparing younger cohorts (BHS+SOL) vs. older cohorts (ARIC+MESA+CHS)

**Table S7.** Associations of polygenic scores (PGS) for sleep traits and global cognitive outcomes in each cohort

**Table S8**. Meta-analyzed effect Estimates and heterogeneity test statistics for the associations of polygenic scores (PGS) for sleep traits and global cognitive outcomes in females and males

**Table S9.** Sex specific associations of polygenic scores (PGS) for sleep traits and global cognitive outcomes in each cohort

**Table S10.** Meta-analyzed effect estimates and heterogeneity test statistics for the associations of polygenic scores (PGS) for sleep traits and global cognitive outcomes in White, Black and Hispanic individuals

**Table S11**. Race/ethnicity specific associations of polygenic scores (PGS) for sleep traits and global cognitive outcomes in each cohort

**Figure S1.** Derivation of global cognitive function (GCF) measures in the study

**Appendix References**

**Appendix 1.** Cohort details

The **Atherosclerosis in Communities (ARIC)** study is a population-based prospective cohort study to study risk factors for subclinical atherosclerosis including 15,792 individuals, predominantly European Americans and African Americans, aged 45-64 years at baseline (1987-89), chosen by probability sampling from four US communities (northwestern suburbs of Minneapolis, Minnesota; Washington County, Maryland; Forsyth County, North Carolina; and Jackson, Mississippi). Cohort members completed three additional triennial follow-up examinations, a fifth exam in 2011-2013, a sixth exam in 2016-2017, a seventh exam in 2018-2019, an eighth exam in 2020, a ninth exam in 2021-2022, a tenth exam in 2023, and an eleventh exam in 2024-2025. A twelfth exam is ongoing. Participants are also contacted by telephone twice a year to report on their health and any hospitalizations. Further details of the cohort design and participants characteristics have been published previously.**^1^**

The **Bogalusa Heart Study (BHS)** is longitudinal population-based study of the semi-rural community in Bogalusa, Louisiana covering both White and Black individuals. Individuals have been followed from childhood to adulthood on roughly a biannual basis for over 40 years and evaluations are still ongoing. Recruitment started January 1, 1972 and ended January 1, 2016. A total of 1298 participants (during their midlife) took part in the first cognitive test visits between 2013 and 2016. Later during 2019-2024, some participated in second cognitive test visits. Further details can be referred to previous publication.^2^

The **Cardiovascular Health Study (CHS)** recruited 5,888 men and women aged 65 or older in four U.S. communities (Sacramento, CA; Hagerstown, MD; Winston-Salem, NC; and Pittsburgh, PA) with annual clinical exams conducted between 1989 and 1999, and twice yearly telephone contacts since 2000. Initial recruitment was in 1989–1990 and an additional cohort of Black participants was added later in 1992–1993. Participants underwent comprehensive baseline assessments including clinical examinations, laboratory testing, imaging, and detailed evaluations of lifestyle and medical history, and were followed longitudinally through regular clinic visits and telephone contacts. Cardiovascular events, hospitalizations, and mortality were systematically ascertained and adjudicated, enabling long-term investigation of incident cardiovascular disease and related outcomes in aging populations. Further details can be referred to previous publication.^3^

The **Multi-Ethnic Study of Atherosclerosis (MESA)** is a study of the characteristics of subclinical cardiovascular disease and the risk factors that predict progression to clinically overt cardiovascular disease or progression of the subclinical disease.**^4^** MESA consists of a diverse, community-based sample of an initial 6,814 men and women aged 45-84 years without known cardiovascular disease at baseline. Thirty-eight percent of the recruited participants were White, 28 percent African American, 22 percent Hispanic, and 12 percent of Chinese descent. Participants were recruited from six fi eld centers across the United States: Baltimore City and Baltimore County, Maryland; Chicago, Illinois; Forsyth County, North Carolina; Los Angeles County, California; New York, New York; and St. Paul, Minnesota. The first examination took place over two years, from July 2000 to July 2002,and has been followed by additional examinations.

Participants are being followed for identification and characterization of cardiovascular disease events, including acute myocardial infarction and other forms of coronary heart disease (CHD), stroke, and heart failure; for cardiovascular disease interventions; and for mortality. Follow-up telephone interviews with participants or their proxies are attempted at least annually to identify new hospitalizations and diagnoses. MESA staff request information on hospital admissions for any reason, outpatient CVD diagnoses, and death. Death certificate records are obtained from state vital statistics or the National Death Index. MESA staff record International Classification of Disease (ICD) diagnosis codes for all hospitalizations and request medical records for those with ICD codes related to CVD. MESA staff also request outpatient records for potential CVD events. However, not all hospitalizations and/or CVD event records were fully documented. Participants underwent clinical examinations covering laboratory tests, imaging, and multiple physical health outcomes at each visit, as well as participated in sleep questionnaires (visit 5) in the MESA Sleep Ancillary Study**^5^**, and cognitive function exams (visits 5-7).

The study was approved by the Institutional review boards at all participating institutions, and all participants gave written informed consent. In addition, informed consent was obtained for extensive data sharing (dbGaP) and genetic/omic studies, including candidate genes (NHLBI CARe), genome-wide scans (NHLBI SHARe), exome sequencing(NHLBI ESP) and, most recently, the NHLBI TOPMed program.

The **Hispanic Community Health Study/Study of Latinos (HCHS/SOL or SOL)** is a longitudinal cohort study covering a total of 16,415 individuals aged 18 to 74 years of Hispanic/Latino origin (Cuban, Puerto Rican, Dominican, Mexican, and Central/South American) at baseline (2008-2011). Individuals from four study centers (Chicago, IL; Miami, FL; Bronx, NY; San Diego, CA) were recruited through a stratified multi-stage area probability sampling design process. Participants went through physical exams and completed multiple questionnaires at baseline which included neurocognitive function tests for a subset of individuals aged 45–74 years. Additional neurocognitive function tests were administered later as part of the Study of Latinos – Investigation of Neurocognitive Aging (SOL-INCA) ancillary study^6^ which occurred at or after the second HCHS/SOL exam (average of 7 years post-baseline visit). Further details can be seen in previous publications on the study design and implementation of HCHS/SOL, and SOL-INCA.^6-8^

**Appendix 2.** Genetic data

**In ARIC,** Genotyping was performed using the Affymetrix GeneChip SNP Array 6.0.^9^ Imputation was subsequently carried out using the TOPMed 2.0 reference panel, with variants having an imputation quality score R² ≤ 0.40 or minor allele frequency (MAF) < 0.01 after imputation being discarded. Sample-level quality control (QC) removed individuals with sex discrepancies, first-degree relatives of included participants, genetic outliers identified through allele sharing and principal component (PC) analyses, and those with a call rate below 95%. We excluded single nucleotide polymorphisms (SNPs) with MAF less than 0.01, and those deviating from Hardy-Weinberg equilibrium (HWE) with p < 1x10⁻⁵. Genomic ancestry principal components were calculated by the ‘EIGENSOFT’ software.^10,11^ The first 10 genetic PCs were adjusted in the outcome regression models.

**In BHS,** genome-wide genotyping was conducted using Illumina Human610 BeadChip described previously.^12-15^ Briefly, sample-level QC removed individuals with sex discrepancies and a call rate < 95%. Variant-level QC removed markers with a call rate <95%, MAF < 0.01, or deviation from HWE P < 1 × 10-6. Genotyping were subsequently imputed to the TOPMed 2.0 reference panel using the TOPMed Imputation Server^16-18^, variants with an imputation quality score R^2^ <0.30 were further removed. Genomic ancestry principal components were calculated by the ‘GCTA’ software for White and Black participants separately.^19^ The first 10 genetic PCs were adjusted in the outcome regression models.

**For CHS and MESA,** whole genome sequencing (WGS) genetic data from the NHLBI Trans-Omics for Precision Medicine (TOPMed) Freeze 10b was utilized. Further details on sequencing procedures and quality control can be found on the TOPMed website: https://topmed.nhlbi.nih.gov/data-resources/methods. Genetic PCs (first 10) were calculated previously for MESA using the ‘GENESIS’ R package^20^ following the University of Washington Genetic Analysis Center (UW-GAC) TOPMed analysis pipeline: https://github.com/UW-GAC/analysis_pipeline. For CHS, genetic PCs (first 11) were created from the TOPMed program, and we downloaded them from dbGaP.

**For SOL,** first, WGS data were generated for 13,025 HCHS/SOL participants through the TOPMed program and the Centers for Common Disease Genomics (CCDG) initiative. Sequencing was performed at the Baylor College of Medicine Human Genome Sequencing Center in accordance with TOPMed protocols, and variant calling was performed using Illumina’s DRAGEN pipeline.^21^ Sequence data were aligned to the GRCh38 (hg38) reference genome. For a subset of 1,083 individuals for which WGS was not available, array genotyping and imputed data was used. For the imputed data, genotyping was conducted with the Illumina HumanOmni2.5 8 BeadChip. Further details in terms of genotyping in SOL can be found in previous publication.^22^ Standard quality control procedures included evaluation of call rates, relatedness, population substructure, and batch effects. Imputation was performed using the TOPMed 3.0 reference panel. The WGS and imputed genotypes datasets were then aggregated by (1) filtering the WGS dataset to keep SNPs with MAF of at least 0.01; (2) filtering the imputed genotypes dataset to keep SNPs with imputation quality R^2^ ≥ 0.8 and kept if they overlap with the WGS SNPs. Genetic PCs and kinship estimates for the combined WGS and imputed genotypes data were computed using the "LD Pruning" app, KING --ibdseg^23,24^, and PC-AiR^25^, all as implemented in the UW-GAC Ancestry and Relatedness toolkit on the BioData Catalyst platform. We accounted for the first 5 genetic PCs in the outcome regression models.

**Appendix 3.** Additional discussion

Overall, we found no modification by sex for the observed associations of long sleep and OSA PGSs with global cognitive function at baseline, indicating the observed overall associations are likely not sex-heterogeneous. One exception is that higher OSA PGS was more strongly associated with increased incident risk of poor GCF at the follow-up among females than in males, yet with strong association heterogeneity observed across the cohorts. Sex differences in the pathophysiology, symptoms, and recognition of OSA are well-recognized.^26^ Prior data from the UK Biobank suggests that women with diagnosed OSA are more likely to show selected memory deficits than males with OSA.^27^ MRI imaging studies also have suggested sex differences in the neuropathologic associations with OSA, with OSA associated with larger hippocampal volumes in males but smaller volumes in females, consistent with different vulnerabilities.^28^ However, interpretation of sex difference with incident risk needs to be interpreted cautiously given the strong association heterogeneity observed across the cohorts, limited sample size and small incident case prevalence within each sex group. Our analyses presented a potential moderate modification by race/ethnicity on the association strengths between sleep PGS and global cognition. For example, the associations for long sleep and OSA PGS (after accounting for BMI) with cognition were observed only among White and Hispanic individuals, but not Black individuals. This observed heterogeneity may be due to several considerations. First, the PGSs we developed for sleep traits (except for OSA) used White/European ancestry populations of European ancestries in the original GWAS studies, and these PGS may not generalize well to Black populations due to differences in their genetic architecture (having high proportion of African continental ancestry) including causal allele frequencies, allelic effect sizes, and LD patterns.^29^ Hispanic individuals studied here are a admixture of Native American populations with European and, to a limited extent, African populations, and often share significant genetic similarities with populations of European descent.^22^ Second, the meta-analysis sample size for the Black race/ethnicity group was small (N = 3,174 for baseline GCF and poor GCF; N = 1,370 for GCF change; N = 1,143 for incident GCF at follow-up), limiting statistical power. Third, there could be differential population prevalences of sleep traits by race/ethnicity, Black people are in general more likely to report shorter sleep duration than White people^30,31^ and may be less likely to be diagnosed with insomnia or OSA than White individuals.^32,33^ These differences and biases may further influence the prediction power and distribution patterns of the related sleep PGSs. In addition, moderate association heterogeneity across cohorts was observed for some findings (e.g., BMI adjusted OSA PGS with incident poor GCF, insomnia with GCF change), which may be partially explained by different distributions of demographic characteristics, risk factors and prevalences of sleep phenotypes and genetic profiles across cohorts. Potential modification by age is likely, which may be particularly relevant for the study of GCF change over follow-up since cognitive decline is often more observable among older adults.^34^ In the meta-analysis of older vs. younger cohorts, we additionally find that higher genetic risk for insomnia was associated with cognitive decline among the older cohorts (ARIC, MESA and CHS, average age at first selected visit >60 yrs.), which was consistent with current literature findings on insomnia sleep trait.^35-37^

**Table S1. Cognitive tests used to derive global cognitive outcomes in each cohort**

| **Cognitive Test *** | **Measuring Domain** | **SOL** | **MESA** | **ARIC** | **BHS** | **CHS** |
| --- | --- | --- | --- | --- | --- | --- |
| Six-item Screener ^38^ | orientation and memory | X |  |  |  |  |
| Digit Symbol Substitution test ^39^ | processing speed and executive functions, including attention, working memory, and visual-motor coordination | X | X | X | X | X |
| Seven-Item Verbal Learning test – immediate learning ^40^ | short-term verbal learning and memory | X |  |  |  |  |
| Seven-Item Verbal Learning test – recall ^40^ | long-term verbal recall and recognition | X |  |  |  |  |
| Word Fluency test ^41,42^ | verbal ability, executive functions including working memory, inhibition and cognitive flexibility | X |  | X | X |  |
| Cognitive Abilities Screening Instrument ^43^ | long-term memory, short-term memory, attention, mental concentration, orientation, abstraction/judgment, language abilities, visual construction, and list-generating fluency |  | X |  |  |  |
| Digit Span test ^44^ | verbal short-term memory, working memory |  | X |  | X |  |
| Delayed Word Recall test ^45^ | short-term and long-term memory recall |  |  | X | X |  |
| Logical memory I, II ^46^ | episodic memory and auditory memory |  |  |  | X |  |
| II-Recognition ^47^ | episodic recognition memory, often with added contributions from associative memory and attention |  |  |  | X |  |
| Trail making tests A, B ^48^ | processing speed and visuomotor abilities, executive functions |  |  |  | X |  |
| Modified Mini-Mental State Examination ^49^ | global cognitive function, including orientation, attention, language, and visuospatial abilities |  |  |  |  | X |
| Benton Visual Retention Test ^50^ | visual perception, memory, and visuo-constructive abilities |  |  |  |  | X |

^*^ Available from: ARIC - Exam 4 and 5; BHS – Cognitive Visit 1 and 2; CHS – Year 6 and 11; MESA - Visit 5 and 6; SOL - Visit 1 and 2. ARIC, Atherosclerosis Risk in Communities Study; BHS, Bogalusa Heart Study; CHS, Cardiovascular Health Study; MESA, Multi-Ethnic Study of Atherosclerosis; SOL: Hispanic Community Health Study/Study of Latinos. Please note each test can have slight variation in application across different cohorts. Details of the cognitive function tests used within each cohort can be referred to each study (ARIC^51^, BHS^52,53^, CHS^54^, MESA^55,56^, SOL^40^).

**Table S2. GWAS studies used for the development of polygenic scores (PGS) for sleep traits**

| **Sleep trait** | **Definition** | **Year & author** | **Sample size** | **Population** | **Cohort** | **PMID** |
| --- | --- | --- | --- | --- | --- | --- |
| Insomnia | “Do you have trouble falling asleep at night or do you wake up in the middle of the night over past 4 weeks?” with responses of “never/rarely” (control), “sometimes” (control), “usually” (case), or “prefer not to answer” (missing). | Jansen, et al. 2019 | 1,331,010 | European | UK Biobank | 30804565 |
| OSA | Diagnoses from electronic health records | Kurniansyah, et al. 2025 * | over 1.2 million | Mixed | Million Veteran Program, FinnGen | 40472801 |
| Long Sleep | ≥9 h per night | Dashti, et al. 2019 | 446,118 | European | UK Biobank | 30846698 |
| Short Sleep | <7 h per night | Dashti, et al. 2019 | 446,118 | European | UK Biobank | 30846698 |
| Sleep Duration | Self-reported sleep duration supported by accelerometer-derived estimates | Dashti, et al. 2019 | 446,118 | European | UK Biobank | 30846698 |
| Excessive Daytime Sleepiness | “How likely are you to dose off or fall asleep during the daytime when you don’t mean to?” with response of “Never/rarely” (1), “sometimes” (2), “often” (3), “all of the time” (4), “do not know” (missing), and “prefer not to answer” (missing). | Wang, et al. 2019 | 452,071 | European | UK Biobank | 31409809 |

^*^ Study reporting the development of the OSA PGSs.

**Table S3. Pearson correlations of polygenic scores for sleep traits in each cohort.** Bolded entries indicate high correlation (correlation coefficient > 0.70 or < -0.70).

| **Cohort** | **ARIC** | | | | | | |
| --- | --- | --- | --- | --- | --- | --- | --- |
| **PGS Traits** | Insomnia | Long Sleep | Short Sleep | BMI-adjusted OSA | BMI-unadjusted OSA | Sleep Duration | Daytime Sleepiness |
| Insomnia | 1.0000 | 0.0101 | 0.2893 | 0.3415 | 0.2163 | -0.2984 | 0.0826 |
| Long Sleep | 0.0101 | **1.0000** | 0.0315 | 0.0191 | 0.0084 | 0.1579 | 0.0383 |
| Short Sleep | 0.2893 | 0.0315 | **1.0000** | 0.4356 | 0.2774 | -0.5336 | 0.1076 |
| BMI adjusted OSA | 0.3415 | 0.0191 | 0.4356 | **1.0000** | 0.6947 | -0.4993 | 0.1022 |
| BMI unadjusted OSA | 0.2163 | 0.0084 | 0.2774 | 0.6947 | **1.0000** | -0.3205 | 0.0421 |
| Sleep Duration | -0.2984 | 0.1579 | -0.5336 | -0.4993 | -0.3205 | **1.0000** | -0.0761 |
| Daytime Sleepiness | 0.0826 | 0.0383 | 0.1076 | 0.1022 | 0.0421 | -0.0761 | **1.0000** |
| **Cohort** | **BHS** | | | | | | |
| **PGS Traits** | Insomnia | Long Sleep | Short Sleep | BMI-adjusted OSA | BMI-unadjusted OSA | Sleep Duration | Daytime Sleepiness |
| Insomnia | **1.0000** | 0.2714 | **0.7036** | -0.6634 | -0.6337 | -0.5652 | 0.2897 |
| Long Sleep | 0.2714 | **1.0000** | 0.2614 | -0.3052 | -0.2941 | 0.1451 | 0.2112 |
| Short Sleep | **0.7036** | 0.2614 | **1.0000** | **-0.7435** | **-0.7113** | **-0.8148** | 0.2479 |
| BMI adjusted OSA | -0.6634 | -0.3052 | **-0.7435** | **1.0000** | **0.9711** | 0.5751 | -0.2445 |
| BMI unadjusted OSA | -0.6337 | -0.2941 | **-0.7113** | **0.9711** | **1.0000** | 0.5511 | -0.2047 |
| Sleep Duration | -0.5652 | 0.1451 | **-0.8148** | 0.5751 | 0.5511 | **1.0000** | -0.1370 |
| Daytime Sleepiness | 0.2897 | 0.2112 | 0.2479 | -0.2445 | -0.2047 | -0.1370 | **1.0000** |
| **Cohort** | **CHS** | | | | | | |
| **PGS Traits** | Insomnia | Long Sleep | Short Sleep | BMI adjusted OSA | BMI unadjusted OSA | Sleep Duration | Daytime Sleepiness |
| Insomnia | **1.0000** | 0.1302 | 0.6358 | -0.5586 | -0.5179 | -0.4851 | 0.2259 |
| Long Sleep | 0.1302 | **1.0000** | 0.1360 | -0.1994 | -0.1755 | 0.3226 | 0.1098 |
| Short Sleep | 0.6358 | 0.1360 | **1.0000** | -0.6337 | -0.5945 | **-0.7704** | 0.1801 |
| BMI adjusted OSA | -0.5586 | -0.1994 | -0.6337 | **1.0000** | **0.9525** | 0.4108 | -0.1688 |
| BMI unadjusted OSA | -0.5179 | -0.1755 | -0.5945 | **0.9525** | **1.0000** | 0.3885 | -0.1479 |
| Sleep Duration | -0.4851 | 0.3226 | **-0.7704** | 0.4108 | 0.3885 | **1.0000** | -0.0874 |
| Daytime Sleepiness | 0.2259 | 0.1098 | 0.1801 | -0.1688 | -0.1479 | -0.0874 | **1.0000** |
| **Cohort** | **MESA** | | | | | | |
| **PGS Traits** | Insomnia | Long Sleep | Short Sleep | BMI adjusted OSA | BMI unadjusted OSA | Sleep Duration | Daytime Sleepiness |
| Insomnia | **1.0000** | 0.1805 | 0.6070 | -0.5579 | -0.5271 | -0.4148 | 0.2173 |
| Long Sleep | 0.1805 | **1.0000** | 0.1364 | -0.2626 | -0.2422 | 0.3329 | 0.0970 |
| Short Sleep | 0.6070 | 0.1364 | **1.0000** | -0.5678 | -0.4962 | **-0.7386** | 0.2683 |
| BMI adjusted OSA | -0.5579 | -0.2626 | -0.5678 | **1.0000** | **0.9370** | 0.2712 | -0.1220 |
| BMI unadjusted OSA | -0.5271 | -0.2422 | -0.4962 | **0.9370** | **1.0000** | 0.2205 | -0.0658 |
| Sleep Duration | -0.4148 | 0.3329 | **-0.7386** | 0.2712 | 0.2205 | **1.0000** | -0.1829 |
| Daytime Sleepiness | 0.2173 | 0.0970 | 0.2683 | -0.1220 | -0.0658 | -0.1829 | **1.0000** |
| **Cohort** | **SOL** | | | | | | |
| **PGS Traits** | Insomnia | Long Sleep | Short Sleep | BMI adjusted OSA | BMI unadjusted OSA | Sleep Duration | Daytime Sleepiness |
| Insomnia | **1.0000** | 0.0458 | 0.4114 | -0.1390 | -0.0858 | -0.3283 | 0.1506 |
| Long Sleep | 0.0458 | **1.0000** | 0.0241 | -0.0329 | -0.0178 | 0.4345 | 0.0848 |
| Short Sleep | 0.4114 | 0.0241 | **1.0000** | -0.2359 | -0.1719 | **-0.7063** | 0.1055 |
| BMI adjusted OSA | -0.1390 | -0.0329 | -0.2359 | **1.0000** | **0.9196** | 0.0465 | 0.0947 |
| BMI unadjusted OSA | -0.0858 | -0.0178 | -0.1719 | **0.9196** | **1.0000** | 0.0032 | 0.1333 |
| Sleep Duration | -0.3283 | 0.4345 | **-0.7063** | 0.0465 | 0.0032 | **1.0000** | -0.0675 |
| Daytime Sleepiness | 0.1506 | 0.0848 | 0.1055 | 0.0947 | 0.1333 | -0.0675 | **1.0000** |

**Table S4. Meta-analyzed effect estimates and heterogeneity test statistics for the associations of polygenic scores (PGS) for sleep traits and global cognitive outcomes in overall population.** Estimates are presented as differences in levels or odds ratio (OR) or incident risk ratio (IRR) and their 95% confidence interval (95% CI) per 1 standard deviation increase in each sleep trait PGS accounting for age, sex, race/ethnicity background, study locations, genetic PCs and education at baseline. For GCF change, the years interval between baseline and follow-up visit was additionally adjusted. For incident poor GCF, years interval between baseline and follow-up visit was fitted as an offset. Fixed-effects meta-analysis was applied using effect estimates from ARIC, BHS, CHS, MESA and SOL. Bolded entries indicate significant associations at z test p<0.05 or marginally significant associations at z test p<0.10.

| **Sleep PGS** | **GCF** | | | | **Poor GCF** | | | |
| --- | --- | --- | --- | --- | --- | --- | --- | --- |
|  | **N = 22,685** | | | | **N = 22,685** | | | |
|  | **Estimate (95% CI)** | **Z test *P*** | **Q test *P*** | **I^2^** | **OR (95% CI)** | **Z test *P*** | **Q test *P*** | **I^2^** |
| Insomnia | 0.00 (-0.02, 0.01) | 0.90 | 0.98 | 0 | 1.00 (0.95, 1.06) | 0.90 | 0.54 | 0 |
| Long Sleep | **-0.02 (-0.03, 0.00)** | **0.01** | 0.90 | 0 | **1.04 (1.00, 1.09)** | **0.06** | 0.49 | 0 |
| Short Sleep | 0.00 (-0.01, 0.02) | 0.65 | 0.26 | 24.25 | 0.97 (0.92, 1.02) | 0.23 | 0.54 | 0 |
| Sleep Duration | 0.00 (-0.02, 0.01) | 0.54 | 0.50 | 0 | 1.03 (0.98, 1.09) | 0.19 | 0.87 | 0 |
| Daytime Sleepiness | 0.00 (-0.01, 0.01) | 0.97 | 0.68 | 0 | 0.99 (0.95, 1.04) | 0.72 | 0.59 | 0 |
| BMI adjusted OSA | -0.02 (-0.05, 0.01) | 0.16 | 0.71 | 0 | **1.11 (1.00, 1.22)** | **0.04** | 0.86 | 0 |
| BMI unadjusted OSA | 0.00 (-0.02, 0.02) | 0.80 | 0.15 | 40.89 | 1.01 (0.95, 1.08) | 0.78 | 0.67 | 0 |
| **Sleep PGS** | **GCF Change** | | | | **Incident Poor GCF** | | | |
|  | **N = 11,896** | | | | **N = 10,451** | | | |
|  | **Estimate (95% CI)** | **Z test *P*** | **Q test *P*** | **I^2^** | **IRR (95% CI)** | **Z test *P*** | **Q test *P*** | **I^2^** |
| Insomnia | -0.01 (-0.03, 0.01) | 0.31 | 0.07 | 53.47 | 0.99 (0.92, 1.06) | 0.76 | 0.95 | 0 |
| Long Sleep | 0.00 (-0.02, 0.01) | 0.58 | 0.37 | 6.77 | 1.04 (0.97, 1.11) | 0.27 | 0.67 | 0 |
| Short Sleep | -0.01 (-0.03, 0.01) | 0.27 | 0.46 | 0 | 1.01 (0.94, 1.09) | 0.72 | 0.39 | 3.55 |
| Sleep Duration | 0.01 (-0.01, 0.03) | 0.33 | 0.46 | 0 | 1.02 (0.95, 1.09) | 0.65 | 0.88 | 0 |
| Daytime Sleepiness | 0.00 (-0.02, 0.02) | 0.99 | 0.87 | 0 | 0.98 (0.92, 1.04) | 0.53 | 0.19 | 35.27 |
| BMI adjusted OSA | 0.00 (-0.03, 0.04) | 0.83 | 0.63 | 0 | **1.15 (0.99, 1.33)** | **0.07** | 0.12 | 44.57 |
| BMI unadjusted OSA | 0.01 (-0.02, 0.03) | 0.50 | 0.79 | 0 | 1.06 (0.96, 1.16) | 0.26 | 0.41 | 0 |

**Table S5. Meta-analyzed effect estimates and heterogeneity test statistics for the associations of polygenic scores (PGS) for sleep Traits and global cognitive outcomes in overall population additionally adjusting for APOE e4 allele status.** Estimates are presented as differences in levels or odds ratio (OR) or incident risk ratio (IRR) and their 95% confidence interval (95% CI) per 1 standard deviation increase in each sleep trait PGS accounting for age, sex, race/ethnicity background, study locations, genetic PCs, education at baseline and APOE 4 allele status (0, 1 or 2 copies of APOE 4 allele). For GCF change, the years interval between baseline and follow-up visit was additionally adjusted. For incident poor GCF, years interval between baseline and follow-up visit was fitted as an offset. Fixed-effects meta-analysis was applied using effect estimates from ARIC, BHS, CHS, MESA and SOL. Bolded entries indicate significant associations at z test p<0.05 or marginally significant associations at z test p<0.10.

| **Sleep PGS** | **GCF** | | | | **Poor GCF** | | | |
| --- | --- | --- | --- | --- | --- | --- | --- | --- |
|  | **N = 20,715** | | | | **N = 20,715** | | | |
|  | **Estimate (95% CI)** | **Z test *P*** | **Q test *P*** | **I^2^** | **OR (95% CI)** | **Z test *P*** | **Q test *P*** | **I^2^** |
| Insomnia | 0.00 (-0.02, 0.02) | 0.95 | 0.86 | 0 | 1.01 (0.95, 1.06) | 0.80 | 0.76 | 0 |
| Long Sleep | **-0.02 (-0.03, -0.01)** | **<0.01** | 0.81 | 0 | **1.05 (1.01, 1.10)** | **0.03** | 0.61 | 0 |
| Short Sleep | 0.01 (-0.01, 0.02) | 0.53 | 0.13 | 43.54 | 0.96 (0.91, 1.01) | 0.15 | 0.63 | 0 |
| Sleep Duration | 0.00 (-0.02, 0.01) | 0.69 | 0.23 | 28.37 | 1.03 (0.98, 1.08) | 0.27 | 0.89 | 0 |
| Daytime Sleepiness | 0.00 (-0.01, 0.02) | 0.65 | 0.60 | 0 | 0.99 (0.95, 1.04) | 0.70 | 0.32 | 14.45 |
| BMI adjusted OSA | -0.02 (-0.05, 0.01) | 0.27 | 0.68 | 0 | **1.10 (1.00, 1.22)** | **0.05** | 0.68 | 0 |
| BMI unadjusted OSA | 0.00 (-0.02, 0.02) | 0.94 | 0.22 | 30.16 | 1.00 (0.94, 1.07) | 0.90 | 0.77 | 0 |
| **Sleep PGS** | **GCF Change** | | | | **Incident Poor GCF** | | | |
|  | **N = 11,088** | | | | **N = 9,741** | | | |
|  | **Estimate (95% CI)** | **Z test *P*** | **Q test *P*** | **I^2^** | **IRR (95% CI)** | **Z test *P*** | **Q test *P*** | **I^2^** |
| Insomnia | -0.01 (-0.03, 0.01) | 0.18 | 0.02 | 64.71* | 0.98 (0.91, 1.06) | 0.65 | 0.98 | 0 |
| Long Sleep | -0.01 (-0.02, 0.01) | 0.45 | 0.37 | 5.76 | 1.05 (0.99, 1.13) | 0.13 | 0.60 | 0 |
| Short Sleep | -0.01 (-0.02, 0.01) | 0.43 | 0.72 | 0 | 1.01 (0.94, 1.09) | 0.82 | 0.40 | 1.18 |
| Sleep Duration | 0.00 (-0.01, 0.02) | 0.59 | 0.64 | 0 | 1.03 (0.96, 1.11) | 0.38 | 0.92 | 0 |
| Daytime Sleepiness | 0.00 (-0.02, 0.01) | 0.83 | 0.89 | 0 | 0.98 (0.92, 1.05) | 0.57 | 0.42 | 0 |
| BMI adjusted OSA | 0.00 (-0.04, 0.03) | 0.85 | 0.68 | 0 | **1.15 (0.99, 1.33)** | **0.07** | 0.19 | 35.39 |
| BMI unadjusted OSA | 0.00 (-0.02, 0.03) | 0.74 | 0.48 | 0 | 1.06 (0.96, 1.16) | 0.27 | 0.42 | 0 |

*I-squared is statistically significant (at Q test p<0.05), indicating potential effect heterogeneity across the meta-analysis cohorts.

**Table S6. Meta-analyzed effect estimates and heterogeneity test statistics for the associations of polygenic scores (PGS) for sleep traits and global cognitive outcomes in overall population comparing younger cohorts (BHS+SOL) vs. older cohorts (ARIC+MESA+CHS).** Estimates are presented as differences in levels or odds ratio (OR) or incident risk ratio (IRR) and their 95% confidence interval (95% CI) per 1 standard deviation increase in each sleep trait PGS accounting for age, sex, race/ethnicity background, study locations, genetic PCs and education at baseline. For GCF change, the years interval between baseline and follow-up visit was additionally adjusted. For incident poor GCF, years interval between baseline and follow-up visit was fitted as an offset. Fixed-effects meta-analysis was applied using effect estimates from ARIC, BHS, CHS, MESA and SOL. Bolded entries indicate significant associations at z test p<0.05 or marginally significant associations at z test p<0.10.

| **Sleep PGS** | **Group** | **GCF** | | | | | **Poor GCF** | | | | |
| --- | --- | --- | --- | --- | --- | --- | --- | --- | --- | --- | --- |
|  |  | **N** | **Estimate (95% CI)** | **Z test *P*** | **Q test *P*** | **I^2^** | **N** | **OR (95% CI)** | **Z test *P*** | **Q test *P*** | **I^2^** |
| Insomnia | BHS+SOL | 8,116 | 0 (-0.04, 0.03) | 0.84 | 0.64 | 0.00 | 8,116 | 1.04 (0.94, 1.16) | 0.46 | 0.46 | 0.00 |
|  | ARIC+MESA+CHS | 14,569 | 0 (-0.02, 0.02) | 0.97 | 0.90 | 0.00 | 14,569 | 0.99 (0.94, 1.05) | 0.80 | 0.37 | 0.00 |
| Long Sleep | BHS+SOL | 8,116 | **-0.03 (-0.05, 0)** | **0.04** | 1.00 | 0.00 | 8,116 | **1.11 (1.01, 1.22)** | **0.02** | 0.59 | 0.00 |
|  | ARIC+MESA+CHS | 14,569 | -0.01 (-0.03, 0) | 0.11 | 1.00 | 0.00 | 14,569 | 1.02 (0.97, 1.08) | 0.38 | 0.71 | 0.00 |
| Short Sleep | BHS+SOL | 8,116 | -0.01 (-0.04, 0.02) | 0.35 | 0.22 | 33.77 | 8,116 | 1 (0.89, 1.11) | 0.95 | 0.42 | 0.00 |
|  | ARIC+MESA+CHS | 14,569 | 0.01 (-0.01, 0.03) | 0.26 | 0.40 | 0.00 | 14,569 | 0.96 (0.9, 1.02) | 0.18 | 0.34 | 6.61 |
| Sleep Duration | BHS+SOL | 8,116 | 0 (-0.03, 0.03) | 0.92 | 0.58 | 0.00 | 8,116 | 1.05 (0.96, 1.16) | 0.28 | 0.52 | 0.00 |
|  | ARIC+MESA+CHS | 14,569 | -0.01 (-0.02, 0.01) | 0.45 | 0.24 | 28.98 | 14,569 | 1.03 (0.97, 1.09) | 0.37 | 0.73 | 0.00 |
| Daytime Sleepiness | BHS+SOL | 8,116 | -0.01 (-0.04, 0.01) | 0.37 | 0.39 | 0.00 | 8,116 | 1.03 (0.95, 1.12) | 0.49 | 0.58 | 0.00 |
|  | ARIC+MESA+CHS | 14,569 | 0 (-0.01, 0.02) | 0.59 | 0.78 | 0.00 | 14,569 | 0.98 (0.93, 1.03) | 0.41 | 0.48 | 0.00 |
| BMI adjusted OSA | BHS+SOL | 8,116 | -0.04 (-0.1, 0.02) | 0.18 | 0.41 | 0.00 | 8,116 | 1.13 (0.96, 1.34) | 0.15 | 0.83 | 0.00 |
|  | ARIC+MESA+CHS | 14,569 | -0.01 (-0.05, 0.02) | 0.42 | 0.63 | 0.00 | 14,569 | 1.09 (0.97, 1.23) | 0.13 | 0.56 | 0.00 |
| BMI unadjusted OSA | BHS+SOL | 8,116 | **-0.05 (-0.1, 0)** | **0.05** | 0.90 | 0.00 | 8,116 | 1.09 (0.94, 1.26) | 0.25 | 0.75 | 0.00 |
|  | ARIC+MESA+CHS | 14,569 | 0.01 (-0.02, 0.03) | 0.51 | 0.27 | 24.08 | 14,569 | 0.99 (0.92, 1.06) | 0.81 | 0.61 | 0.00 |
| **Sleep PGS** | **Group** | **GCF Change** | | | | | **Incident Poor GCF** | | | | |
|  |  | **N** | **Estimate (95% CI)** | **Z test *P*** | **Q test *P*** | **I^2^** | **N** | **IRR (95% CI)** | **Z test *P*** | **Q test *P*** | **I^2^** |
| Insomnia | BHS+SOL | 5,224 | 0.02 (-0.01, 0.04) | 0.29 | 0.15 | 52.36 | 4,468 | 0.96 (0.83, 1.1) | 0.53 | 0.61 | 0.00 |
|  | ARIC+MESA+CHS | 6,672 | **-0.02 (-0.04, 0)** | **0.05** | 0.29 | 18.23 | 5,983 | 1 (0.92, 1.09) | 0.97 | 0.94 | 0.00 |
| Long Sleep | BHS+SOL | 5,224 | 0.02 (-0.02, 0.06) | 0.31 | 0.14 | 53.74 | 4,468 | 1.02 (0.9, 1.16) | 0.75 | 0.13 | 55.71 |
|  | ARIC+MESA+CHS | 6,672 | -0.01 (-0.03, 0.01) | 0.30 | 0.85 | 0.00 | 5,983 | 1.04 (0.97, 1.13) | 0.27 | 0.98 | 0.00 |
| Short Sleep | BHS+SOL | 5,224 | 0 (-0.03, 0.03) | 0.77 | 0.53 | 0.00 | 4,468 | 1 (0.88, 1.13) | 0.97 | 0.06 | 72.36 |
|  | ARIC+MESA+CHS | 6,672 | -0.02 (-0.04, 0) | 0.12 | 0.39 | 0.00 | 5,983 | 1.02 (0.93, 1.12) | 0.63 | 0.81 | 0.00 |
| Sleep Duration | BHS+SOL | 5,224 | 0.01 (-0.03, 0.04) | 0.74 | 0.37 | 0.00 | 4,468 | 1.06 (0.93, 1.21) | 0.35 | 0.49 | 0.00 |
|  | ARIC+MESA+CHS | 6,672 | 0.01 (-0.01, 0.03) | 0.36 | 0.24 | 29.04 | 5,983 | 1 (0.92, 1.09) | 0.95 | 0.98 | 0.00 |
| Daytime Sleepiness | BHS+SOL | 5,224 | 0.01 (-0.02, 0.04) | 0.69 | 0.95 | 0.00 | 4,468 | 1.01 (0.91, 1.11) | 0.87 | 0.13 | 55.67 |
|  | ARIC+MESA+CHS | 6,672 | 0 (-0.02, 0.02) | 0.83 | 0.60 | 0.00 | 5,983 | 0.96 (0.89, 1.04) | 0.36 | 0.18 | 42.06 |
| BMI adjusted OSA | BHS+SOL | 5,224 | 0.04 (-0.02, 0.1) | 0.22 | 0.87 | 0.00 | 4,468 | 1.08 (0.82, 1.43) | 0.57 | 0.04 | 76.79* |
|  | ARIC+MESA+CHS | 6,672 | -0.01 (-0.05, 0.03) | 0.59 | 0.66 | 0.00 | 5,983 | **1.17 (0.99, 1.39)** | **0.07** | 0.26 | 25.88 |
| BMI unadjusted OSA | BHS+SOL | 5,224 | 0.02 (-0.03, 0.07) | 0.48 | 0.92 | 0.00 | 4,468 | **1.19 (0.98, 1.44)** | **0.08** | 0.21 | 36.87 |
|  | ARIC+MESA+CHS | 6,672 | 0.01 (-0.02, 0.03) | 0.69 | 0.48 | 0.00 | 5,983 | 1.02 (0.91, 1.13) | 0.75 | 0.76 | 0.00 |

*I-squared is statistically significant (at Q test p<0.05), indicating potential effect heterogeneity across the meta-analysis cohorts.

**Table S7. Associations of polygenic scores (PGS) for sleep traits and global cognitive outcomes in each cohort.** Estimates are presented as differences in levels or odds ratio (OR) or incident risk ratio (IRR) and their 95% confidence interval (95% CI) per 1 standard deviation increase in each sleep trait PGS accounting for age, sex, race/ethnicity background, study locations, genetic PCs and education at baseline. For GCF change, the years interval between baseline and follow-up visit was additionally adjusted. For incident poor GCF, years interval between baseline and follow-up visit was fitted as an offset. Bolded entries indicate significant associations at z test p<0.05 or marginally significant associations at z test p<0.10.

| **Sleep PGS** | **Cohort** | **GCF** | | | **Poor GCF** | | |
| --- | --- | --- | --- | --- | --- | --- | --- |
|  |  | **N** | **Estimate** | ***P*** | **N** | **OR** | ***P*** |
|  |  |  | **(95% CI)** |  |  | **(95% CI)** |  |
| Insomnia | ARIC | 8,086 | 0 (-0.02, 0.03) | 0.77 | 8,086 | 0.98 (0.92, 1.06) | 0.66 |
|  | BHS | 1,078 | -0.02 (-0.09, 0.05) | 0.61 | 1,078 | 1.14 (0.88, 1.47) | 0.32 |
|  | CHS | 2,656 | 0 (-0.04, 0.04) | 0.92 | 2,656 | 1.08 (0.94, 1.26) | 0.28 |
|  | MESA | 3,827 | -0.01 (-0.04, 0.03) | 0.73 | 3,827 | 0.94 (0.82, 1.09) | 0.42 |
|  | SOL | 7,038 | 0 (-0.04, 0.04) | 0.96 | 7,038 | 1.02 (0.91, 1.15) | 0.72 |
| Long Sleep | ARIC | 8,086 | -0.01 (-0.04, 0.01) | 0.34 | 8,086 | 1.02 (0.96, 1.09) | 0.54 |
|  | BHS | 1,078 | -0.03 (-0.08, 0.02) | 0.30 | 1,078 | 1.16 (0.96, 1.41) | 0.12 |
|  | CHS | 2,656 | -0.01 (-0.04, 0.02) | 0.41 | 2,656 | 0.99 (0.88, 1.11) | 0.87 |
|  | MESA | 3,827 | -0.01 (-0.04, 0.01) | 0.32 | 3,827 | 1.06 (0.95, 1.18) | 0.31 |
|  | SOL | 7,038 | **-0.03 (-0.06, 0)** | **0.07** | 7,038 | **1.1 (0.99, 1.21)** | **0.08** |
| Short Sleep | ARIC | 8,086 | 0.02 (-0.01, 0.04) | 0.18 | 8,086 | 0.95 (0.88, 1.02) | 0.14 |
|  | BHS | 1,078 | -0.06 (-0.14, 0.02) | 0.14 | 1,078 | 1.11 (0.83, 1.48) | 0.47 |
|  | CHS | 2,656 | 0.02 (-0.02, 0.06) | 0.31 | 2,656 | 1.06 (0.91, 1.24) | 0.43 |
|  | MESA | 3,827 | -0.01 (-0.04, 0.02) | 0.60 | 3,827 | 0.92 (0.79, 1.07) | 0.29 |
|  | SOL | 7,038 | -0.01 (-0.04, 0.02) | 0.68 | 7,038 | 0.98 (0.87, 1.1) | 0.72 |
| Sleep Duration | ARIC | 8,086 | -0.01 (-0.04, 0.02) | 0.54 | 8,086 | 1.03 (0.96, 1.11) | 0.42 |
|  | BHS | 1,078 | 0.02 (-0.04, 0.08) | 0.59 | 1,078 | 0.98 (0.78, 1.24) | 0.90 |
|  | CHS | 2,656 | -0.03 (-0.06, 0.01) | 0.11 | 2,656 | 0.98 (0.86, 1.12) | 0.79 |
|  | MESA | 3,827 | 0.01 (-0.02, 0.04) | 0.50 | 3,827 | 1.05 (0.93, 1.19) | 0.40 |
|  | SOL | 7,038 | 0 (-0.03, 0.03) | 0.86 | 7,038 | 1.07 (0.96, 1.19) | 0.22 |
| Daytime Sleepiness | ARIC | 8,086 | 0 (-0.02, 0.03) | 0.92 | 8,086 | 1 (0.94, 1.07) | 0.97 |
|  | BHS | 1,078 | 0.01 (-0.04, 0.06) | 0.79 | 1,078 | 0.98 (0.81, 1.19) | 0.85 |
|  | CHS | 2,656 | 0 (-0.03, 0.03) | 0.92 | 2,656 | 0.92 (0.82, 1.04) | 0.19 |
|  | MESA | 3,827 | 0.01 (-0.01, 0.04) | 0.39 | 3,827 | 0.96 (0.86, 1.08) | 0.51 |
|  | SOL | 7,038 | -0.02 (-0.05, 0.01) | 0.23 | 7,038 | 1.04 (0.95, 1.15) | 0.39 |
| BMI adjusted OSA | ARIC | 8,086 | -0.01 (-0.06, 0.04) | 0.66 | 8,086 | 1.1 (0.95, 1.26) | 0.19 |
|  | BHS | 1,078 | -0.11 (-0.3, 0.07) | 0.23 | 1,078 | 1.22 (0.61, 2.46) | 0.58 |
|  | CHS | 2,656 | 0.02 (-0.07, 0.1) | 0.69 | 2,656 | 0.94 (0.67, 1.31) | 0.71 |
|  | MESA | 3,827 | -0.03 (-0.09, 0.03) | 0.27 | 3,827 | 1.19 (0.91, 1.55) | 0.21 |
|  | SOL | 7,038 | -0.03 (-0.09, 0.03) | 0.30 | 7,038 | 1.13 (0.95, 1.34) | 0.18 |
| BMI unadjusted OSA | ARIC | 8,086 | 0.02 (-0.01, 0.05) | 0.15 | 8,086 | 0.98 (0.9, 1.06) | 0.55 |
|  | BHS | 1,078 | -0.06 (-0.19, 0.08) | 0.42 | 1,078 | 1.01 (0.61, 1.67) | 0.98 |
|  | CHS | 2,656 | -0.01 (-0.07, 0.06) | 0.87 | 2,656 | 1.02 (0.78, 1.33) | 0.90 |
|  | MESA | 3,827 | -0.02 (-0.07, 0.02) | 0.33 | 3,827 | 1.1 (0.88, 1.37) | 0.41 |
|  | SOL | 7,038 | **-0.05 (-0.1, 0)** | **0.08** | 7,038 | 1.1 (0.94, 1.28) | 0.24 |
| **Sleep PGS** | **Cohort** | **GCF Change** | | | **Incident Poor GCF** | | |
|  |  | **N** | **Estimate** | ***P*** | **N** | **IRR** | ***P*** |
|  |  |  | **(95% CI)** |  |  | **(95% CI)** |  |
| Insomnia | ARIC | 3,530 | -0.01 (-0.04, 0.02) | 0.48 | 3,196 | 1 (0.9, 1.11) | 0.97 |
|  | BHS | 585 | -0.1 (-0.27, 0.06) | 0.22 | 501 | 1 (0.8, 1.25) | 1.00 |
|  | CHS | 1,639 | **-0.05 (-0.08, -0.01)** | **0.02** | 1,435 | 0.98 (0.8, 1.19) | 0.82 |
|  | MESA | 1,503 | -0.01 (-0.05, 0.03) | 0.68 | 1,352 | 1.03 (0.82, 1.3) | 0.79 |
|  | SOL | 4,639 | 0.02 (-0.01, 0.05) | 0.19 | 3,967 | 0.93 (0.78, 1.11) | 0.42 |
| Long Sleep | ARIC | 3,530 | 0 (-0.03, 0.03) | 0.85 | 3,196 | 1.04 (0.94, 1.15) | 0.46 |
|  | BHS | 585 | **0.11 (-0.02, 0.24)** | **0.09** | 501 | 0.92 (0.76, 1.11) | 0.38 |
|  | CHS | 1,639 | -0.01 (-0.04, 0.02) | 0.49 | 1,435 | 1.04 (0.89, 1.22) | 0.60 |
|  | MESA | 1,503 | -0.01 (-0.04, 0.02) | 0.35 | 1,352 | 1.06 (0.89, 1.26) | 0.52 |
|  | SOL | 4,639 | 0.01 (-0.03, 0.05) | 0.61 | 3,967 | 1.12 (0.94, 1.34) | 0.21 |
| Short Sleep | ARIC | 3,530 | -0.02 (-0.05, 0.01) | 0.22 | 3,196 | 1.03 (0.92, 1.15) | 0.58 |
|  | BHS | 585 | 0.06 (-0.12, 0.25) | 0.50 | 501 | 0.81 (0.63, 1.04) | 0.09 |
|  | CHS | 1,639 | -0.03 (-0.07, 0.01) | 0.10 | 1,435 | 1.04 (0.85, 1.27) | 0.69 |
|  | MESA | 1,503 | 0.01 (-0.04, 0.05) | 0.74 | 1,352 | 0.95 (0.74, 1.21) | 0.67 |
|  | SOL | 4,639 | 0 (-0.03, 0.03) | 0.85 | 3,967 | 1.07 (0.93, 1.22) | 0.38 |
| Sleep Duration | ARIC | 3,530 | 0.01 (-0.02, 0.04) | 0.50 | 3,196 | 0.99 (0.89, 1.11) | 0.92 |
|  | BHS | 585 | 0.07 (-0.07, 0.21) | 0.34 | 501 | 1.12 (0.92, 1.37) | 0.26 |
|  | CHS | 1,639 | 0.03 (-0.01, 0.06) | 0.11 | 1,435 | 0.99 (0.84, 1.17) | 0.91 |
|  | MESA | 1,503 | -0.01 (-0.05, 0.02) | 0.45 | 1,352 | 1.01 (0.83, 1.23) | 0.88 |
|  | SOL | 4,639 | 0 (-0.04, 0.04) | 0.92 | 3,967 | 1.02 (0.86, 1.22) | 0.80 |
| Daytime Sleepiness | ARIC | 3,530 | 0 (-0.02, 0.03) | 0.78 | 3,196 | 0.93 (0.84, 1.03) | 0.15 |
|  | BHS | 585 | 0 (-0.12, 0.12) | 0.97 | 501 | 1.12 (0.95, 1.32) | 0.19 |
|  | CHS | 1,639 | 0 (-0.03, 0.03) | 0.82 | 1,435 | 0.94 (0.81, 1.1) | 0.47 |
|  | MESA | 1,503 | -0.02 (-0.05, 0.02) | 0.33 | 1,352 | 1.13 (0.94, 1.37) | 0.19 |
|  | SOL | 4,639 | 0.01 (-0.02, 0.04) | 0.68 | 3,967 | 0.95 (0.84, 1.08) | 0.44 |
| BMI adjusted OSA | ARIC | 3,530 | -0.03 (-0.09, 0.03) | 0.38 | 3,196 | **1.27 (1.03, 1.56)** | **0.02** |
|  | BHS | 585 | 0.08 (-0.37, 0.52) | 0.73 | 501 | **1.74 (1.03, 2.95)** | **0.04** |
|  | CHS | 1,639 | -0.02 (-0.1, 0.07) | 0.72 | 1,435 | 0.84 (0.53, 1.33) | 0.46 |
|  | MESA | 1,503 | 0.02 (-0.06, 0.1) | 0.64 | 1,352 | 1.1 (0.71, 1.69) | 0.67 |
|  | SOL | 4,639 | 0.04 (-0.03, 0.1) | 0.23 | 3,967 | 0.9 (0.65, 1.25) | 0.54 |
| BMI unadjusted OSA | ARIC | 3,530 | 0.02 (-0.02, 0.05) | 0.36 | 3,196 | 1 (0.89, 1.13) | 0.97 |
|  | BHS | 585 | 0 (-0.33, 0.34) | 0.99 | 501 | **1.52 (0.99, 2.33)** | **0.06** |
|  | CHS | 1,639 | -0.03 (-0.1, 0.04) | 0.38 | 1,435 | 1.08 (0.76, 1.54) | 0.65 |
|  | MESA | 1,503 | 0 (-0.06, 0.07) | 0.95 | 1,352 | 1.13 (0.79, 1.6) | 0.51 |
|  | SOL | 4,639 | 0.02 (-0.04, 0.08) | 0.47 | 3,967 | 1.11 (0.9, 1.38) | 0.33 |

**Table S8. Meta-analyzed effect estimates and heterogeneity test statistics for the associations of polygenic scores (PGS) for sleep traits and global cognitive outcomes in females and males.** Estimates are presented as differences in levels or odds ratio (OR) or incident risk ratio (IRR) and their 95% confidence interval (95% CI) per 1 standard deviation increase in each sleep trait PGS accounting for age, sex, race/ethnicity background, study locations, genetic PCs and education at baseline. For GCF change, the years interval between baseline and follow-up visit was additionally adjusted. For incident poor GCF, years interval between baseline and follow-up visit was fitted as an offset. Fixed-effects meta-analysis was applied using effect estimates from ARIC, BHS, CHS, MESA and SOL. Bolded entries indicate significant associations at z test p<0.05 or marginally significant associations at z test p<0.10.

| **Sleep PGS** | **Group** | **GCF** | | | | | **Poor GCF** | | | | |
| --- | --- | --- | --- | --- | --- | --- | --- | --- | --- | --- | --- |
|  |  | **N** | **Estimate (95% CI)** | **Z test *P*** | **Q test *P*** | **I^2^** | **N** | **OR (95% CI)** | **Z test *P*** | **Q test *P*** | **I^2^** |
| Insomnia | Females | 12,965 | -0.01 (-0.03, 0.01) | 0.47 | 0.47 | 0 | 12,965 | 1.01 (0.93, 1.09) | 0.87 | 0.2 | 32.47 |
|  | Males | 9,720 | 0.01 (-0.02, 0.03) | 0.52 | 0.51 | 0 | 9,720 | 1.01 (0.95, 1.08) | 0.73 | 0.8 | 0 |
| Long Sleep | Females | 12,965 | -0.01 (-0.03, 0.00) | 0.11 | 0.94 | 0 | 12,965 | 1.04 (0.97, 1.11) | 0.29 | 0.4 | 0.43 |
|  | Males | 9,720 | **-0.02 (-0.04, 0.00)** | **0.07** | 0.82 | 0 | 9,720 | 1.05 (0.99, 1.11) | 0.13 | 0.59 | 0 |
| Short Sleep | Females | 12,965 | 0.00 (-0.02, 0.03) | 0.66 | 0.25 | 26.24 | 12,965 | **0.93 (0.86, 1.01)** | **0.09** | 0.62 | 0 |
|  | Males | 9,720 | 0.00 (-0.02, 0.02) | 0.98 | 0.75 | 0 | 9,720 | 1.01 (0.94, 1.08) | 0.82 | 0.63 | 0 |
| Sleep Duration | Females | 12,965 | 0.00 (-0.02, 0.02) | 0.77 | 0.29 | 19.25 | 12,965 | 1.05 (0.97, 1.13) | 0.20 | 0.7 | 0 |
|  | Males | 9,720 | 0.00 (-0.02, 0.02) | 0.72 | 0.76 | 0 | 9,720 | 1.01 (0.95, 1.08) | 0.75 | 0.35 | 10.14 |
| Daytime Sleepiness | Females | 12,965 | 0.00 (-0.02, 0.01) | 0.74 | 0.96 | 0 | 12,965 | 0.99 (0.92, 1.06) | 0.70 | 0.72 | 0 |
|  | Males | 9,720 | 0.00 (-0.02, 0.02) | 0.84 | 0.56 | 0 | 9,720 | 0.99 (0.94, 1.06) | 0.85 | 0.69 | 0 |
| BMI adjusted OSA | Females | 12,965 | -0.02 (-0.06, 0.02) | 0.38 | 0.46 | 0 | 12,965 | 1.11 (0.96, 1.29) | 0.16 | 0.57 | 0 |
|  | Males | 9,720 | -0.02 (-0.06, 0.02) | 0.37 | 0.71 | 0 | 9,720 | 1.10 (0.97, 1.24) | 0.14 | 0.71 | 0 |
| BMI unadjusted OSA | Females | 12,965 | 0.00 (-0.03, 0.03) | 1.00 | 0.02 | 64.11* | 12,965 | 1.00 (0.90, 1.11) | 0.99 | 0.51 | 0 |
|  | Males | 9,720 | 0.00 (-0.03, 0.03) | 0.81 | 0.67 | 0 | 9,720 | 1.01 (0.93, 1.10) | 0.83 | 0.64 | 0 |
| **Sleep PGS** | **Group** | **GCF Change** | | | | | **Incident Poor GCF** | | | | |
|  |  | **N** | **Estimate (95% CI)** | **Z test *P*** | **Q test *P*** | **I^2^** | **N** | **IRR (95% CI)** | **Z test *P*** | **Q test *P*** | **I^2^** |
| Insomnia | Females | 7,112 | -0.01 (-0.03, 0.01) | 0.35 | 0.42 | 0 | 7,778 | 0.97 (0.88, 1.06) | 0.51 | 0.03 | 62.76* |
|  | Males | 4,784 | -0.01 (-0.03, 0.02) | 0.64 | 0.08 | 52.58 | 4,032 | 0.92 (0.82, 1.03) | 0.14 | 0.85 | 0 |
| Long Sleep | Females | 7,112 | 0.00 (-0.02, 0.02) | 0.82 | 0.97 | 0 | 7,778 | 1.03 (0.95, 1.12) | 0.47 | 0.06 | 56.23 |
|  | Males | 4,784 | -0.01 (-0.03, 0.01) | 0.34 | 0.23 | 28.46 | 4,032 | 1.03 (0.93, 1.14) | 0.62 | 0.73 | 0 |
| Short Sleep | Females | 7,112 | 0.00 (-0.02, 0.02) | 0.94 | 0.21 | 31.4 | 7,778 | 0.97 (0.89, 1.06) | 0.52 | <0.01 | 76.28* |
|  | Males | 4,784 | -0.02 (-0.04, 0.01) | 0.19 | 0.87 | 0 | 4,032 | 1.01 (0.90, 1.12) | 0.90 | 0.19 | 34.08 |
| Sleep Duration | Females | 7,112 | 0.01 (-0.01, 0.04) | 0.25 | 0.12 | 45.88 | 7,778 | 1.02 (0.93, 1.12) | 0.66 | 0.02 | 66.87* |
|  | Males | 4,784 | -0.01 (-0.03, 0.02) | 0.68 | 0.21 | 31.52 | 4,032 | 1.09 (0.98, 1.21) | 0.13 | 0.87 | 0 |
| Daytime Sleepiness | Females | 7,112 | 0.00 (-0.02, 0.02) | 0.75 | 0.74 | 0 | 7,778 | 0.96 (0.89, 1.03) | 0.26 | 0.57 | 0 |
|  | Males | 4,784 | 0.00 (-0.03, 0.02) | 0.76 | 0.99 | 0 | 4,032 | 0.98 (0.89, 1.07) | 0.60 | 0.76 | 0 |
| BMI adjusted OSA | Females | 7,112 | -0.02 (-0.07, 0.02) | 0.29 | 0.89 | 0 | 7,778 | **1.22 (1.02, 1.45)** | **0.03** | <0.01 | 73.79* |
|  | Males | 4,784 | 0.03 (-0.02, 0.08) | 0.27 | 0.25 | 26.34 | 4,032 | 1.11 (0.91, 1.36) | 0.31 | 0.03 | 62.99* |
| BMI unadjusted OSA | Females | 7,112 | 0.00 (-0.04, 0.03) | 0.89 | 0.85 | 0 | 7,778 | **1.11 (0.99, 1.25)** | **0.08** | <0.01 | 77.12* |
|  | Males | 4,784 | 0.02 (-0.02, 0.05) | 0.32 | 0.88 | 0 | 4,032 | 1.06 (0.93, 1.20) | 0.40 | 0.16 | 38.5 |

*I-squared is statistically significant (at Q test p<0.05), indicating potential effect heterogeneity across the meta-analysis cohorts.

**Table S9. Sex specific associations of polygenic scores (PGS) for sleep traits and global cognitive outcomes in each cohort.** Estimates are presented as differences in levels or odds ratio (OR) or incident risk ratio (IRR) and their 95% confidence interval (95% CI) per 1 standard deviation increase in each sleep trait PGS accounting for age, race/ethnicity background, study locations, genetic PCs and education at baseline. For GCF change, the years interval between baseline and follow-up visit was additionally adjusted. For incident poor GCF, years interval between baseline and follow-up visit was fitted as an offset. Bolded entries indicate significant associations at z test p<0.05 or marginally significant associations at z test p<0.10.

| **Sleep PGS** | **Cohort** | **GCF** | | | | | |
| --- | --- | --- | --- | --- | --- | --- | --- |
|  |  | **Females** | | | **Males** | | |
|  |  | **N** | **Estimate** | ***P*** | **N** | **Estimate** | ***P*** |
|  |  |  | **(95% CI)** |  |  | **(95% CI)** |  |
| Insomnia | ARIC | 4,455 | -0.02 (-0.05,0.02) | 0.34 | 3,631 | 0.03 (-0.01,0.06) | 0.19 |
|  | BHS | 638 | -0.05 (-0.14,0.04) | 0.29 | 440 | 0.03 (-0.08,0.13) | 0.63 |
|  | CHS | 1,574 | -0.02 (-0.07,0.03) | 0.50 | 1,082 | 0.02 (-0.04,0.07) | 0.52 |
|  | MESA | 2,022 | -0.01 (-0.05,0.03) | 0.65 | 1,805 | 0.00 (-0.05,0.04) | 0.90 |
|  | SOL | 4,276 | 0.03 (-0.02,0.07) | 0.24 | 2,762 | -0.03 (-0.09,0.02) | 0.25 |
| Long Sleep | ARIC | 4,455 | -0.01 (-0.04,0.02) | 0.57 | 3,631 | -0.01 (-0.05,0.02) | 0.42 |
|  | BHS | 638 | 0.00 (-0.07,0.07) | 0.96 | 440 | -0.05 (-0.13,0.02) | 0.17 |
|  | CHS | 1,574 | -0.01 (-0.06,0.03) | 0.48 | 1,082 | -0.01 (-0.05,0.04) | 0.81 |
|  | MESA | 2,022 | -0.01 (-0.05,0.02) | 0.44 | 1,805 | -0.01 (-0.04,0.02) | 0.52 |
|  | SOL | 4,276 | -0.03 (-0.07,0.01) | 0.16 | 2,762 | -0.03 (-0.07,0.01) | 0.17 |
| Short Sleep | ARIC | 4,455 | 0.01 (-0.02,0.05) | 0.42 | 3,631 | 0.02 (-0.02,0.06) | 0.30 |
|  | BHS | 638 | **-0.09 (-0.20,0.01)** | **0.07** | 440 | -0.02 (-0.13,0.10) | 0.80 |
|  | CHS | 1,574 | 0.03 (-0.02,0.09) | 0.20 | 1,082 | 0.00 (-0.06,0.06) | 0.97 |
|  | MESA | 2,022 | 0.00 (-0.05,0.05) | 0.98 | 1,805 | -0.02 (-0.07,0.03) | 0.42 |
|  | SOL | 4,276 | -0.01 (-0.05,0.03) | 0.75 | 2,762 | -0.01 (-0.06,0.04) | 0.70 |
| Sleep Duration | ARIC | 4,455 | 0.01 (-0.03,0.04) | 0.70 | 3,631 | -0.02 (-0.06,0.02) | 0.25 |
|  | BHS | 638 | 0.05 (-0.04,0.13) | 0.27 | 440 | -0.02 (-0.11,0.07) | 0.70 |
|  | CHS | 1,574 | **-0.04 (-0.09,0.00)** | **0.06** | 1,082 | 0.00 (-0.05,0.05) | 0.91 |
|  | MESA | 2,022 | 0.01 (-0.03,0.04) | 0.77 | 1,805 | 0.01 (-0.02,0.05) | 0.48 |
|  | SOL | 4,276 | 0.00 (-0.05,0.04) | 0.85 | 2,762 | 0.00 (-0.04,0.04) | 0.95 |
| Daytime Sleepiness | ARIC | 4,455 | -0.01 (-0.04,0.03) | 0.73 | 3,631 | 0.01 (-0.03,0.04) | 0.73 |
|  | BHS | 638 | 0.00 (-0.07,0.07) | 0.97 | 440 | 0.03 (-0.05,0.11) | 0.51 |
|  | CHS | 1,574 | 0.00 (-0.04,0.04) | 0.93 | 1,082 | 0.00 (-0.05,0.05) | 1.00 |
|  | MESA | 2,022 | 0.01 (-0.03,0.04) | 0.73 | 1,805 | 0.02 (-0.02,0.05) | 0.38 |
|  | SOL | 4,276 | -0.02 (-0.06,0.03) | 0.49 | 2,762 | -0.03 (-0.07,0.01) | 0.20 |
| BMI adjusted OSA | ARIC | 4,455 | -0.03 (-0.10,0.04) | 0.43 | 3,631 | 0.02 (-0.06,0.09) | 0.67 |
|  | BHS | 638 | -0.20 (-0.45,0.04) | 0.11 | 440 | 0.02 (-0.27,0.31) | 0.92 |
|  | CHS | 1,574 | 0.04 (-0.08,0.15) | 0.53 | 1,082 | -0.02 (-0.15,0.11) | 0.81 |
|  | MESA | 2,022 | 0.00 (-0.08,0.09) | 0.91 | 1,805 | -0.07 (-0.15,0.02) | 0.13 |
|  | SOL | 4,276 | -0.04 (-0.12,0.05) | 0.38 | 2,762 | -0.03 (-0.10,0.05) | 0.51 |
| BMI unadjusted OSA | ARIC | 4,455 | **0.04 (0.00,0.08)** | **0.05** | 3,631 | 0.00 (-0.04,0.05) | 0.82 |
|  | BHS | 638 | **-0.16 (-0.34,0.02)** | **0.08** | 440 | 0.12 (-0.10,0.33) | 0.29 |
|  | CHS | 1,574 | -0.02 (-0.11,0.07) | 0.64 | 1,082 | 0.02 (-0.09,0.12) | 0.76 |
|  | MESA | 2,022 | -0.02 (-0.09,0.05) | 0.62 | 1,805 | -0.03 (-0.10,0.04) | 0.44 |
|  | SOL | 4,276 | **-0.07 (-0.14,0.00)** | **0.05** | 2,762 | -0.03 (-0.10,0.04) | 0.45 |
| **Sleep PGS** | **Cohort** | **Poor GCF** | | | | | |
|  |  | **Females** | | | **Males** | | |
|  |  | **N** | **OR** | ***P*** | **N** | **OR** | ***P*** |
|  |  |  | **(95% CI)** |  |  | **(95% CI)** |  |
| Insomnia | ARIC | 4,455 | 1.00 (0.89,1.12) | 0.93 | 3,631 | 0.99 (0.90,1.08) | 0.79 |
|  | BHS | 638 | 1.19 (0.83,1.70) | 0.35 | 440 | 1.07 (0.74,1.55) | 0.71 |
|  | CHS | 1,574 | **1.21 (0.99,1.49)** | **0.06** | 1,082 | 0.97 (0.77,1.21) | 0.77 |
|  | MESA | 2,022 | 0.89 (0.73,1.09) | 0.26 | 1,805 | 1.01 (0.82,1.25) | 0.90 |
|  | SOL | 4,276 | 0.96 (0.81,1.13) | 0.59 | 2,762 | 1.10 (0.94,1.28) | 0.23 |
| Long Sleep | ARIC | 4,455 | 0.97 (0.88,1.08) | 0.62 | 3,631 | 1.05 (0.97,1.14) | 0.24 |
|  | BHS | 638 | 1.17 (0.89,1.54) | 0.27 | 440 | 1.16 (0.89,1.52) | 0.28 |
|  | CHS | 1,574 | 1.04 (0.89,1.22) | 0.62 | 1,082 | 0.92 (0.76,1.10) | 0.35 |
|  | MESA | 2,022 | 1.03 (0.88,1.20) | 0.73 | 1,805 | 1.08 (0.92,1.27) | 0.32 |
|  | SOL | 4,276 | **1.16 (0.99,1.37)** | **0.07** | 2,762 | 1.05 (0.93,1.19) | 0.44 |
| Short Sleep | ARIC | 4,455 | **0.90 (0.80,1.01)** | **0.07** | 3,631 | 0.98 (0.90,1.08) | 0.74 |
|  | BHS | 638 | 1.23 (0.82,1.83) | 0.31 | 440 | 1.00 (0.65,1.54) | 0.99 |
|  | CHS | 1,574 | 0.99 (0.81,1.22) | 0.95 | 1,082 | 1.19 (0.93,1.51) | 0.16 |
|  | MESA | 2,022 | 0.91 (0.73,1.13) | 0.38 | 1,805 | 0.94 (0.74,1.18) | 0.58 |
|  | SOL | 4,276 | 0.93 (0.76,1.13) | 0.45 | 2,762 | 1.03 (0.91,1.17) | 0.64 |
| Sleep Duration | ARIC | 4,455 | 1.08 (0.95,1.23) | 0.23 | 3,631 | 0.99 (0.90,1.09) | 0.90 |
|  | BHS | 638 | 0.87 (0.63,1.20) | 0.41 | 440 | 1.11 (0.79,1.54) | 0.55 |
|  | CHS | 1,574 | 1.07 (0.90,1.27) | 0.43 | 1,082 | 0.86 (0.70,1.05) | 0.14 |
|  | MESA | 2,022 | 0.99 (0.83,1.18) | 0.91 | 1,805 | 1.12 (0.93,1.35) | 0.22 |
|  | SOL | 4,276 | 1.10 (0.92,1.32) | 0.31 | 2,762 | 1.04 (0.92,1.19) | 0.52 |
| Daytime Sleepiness | ARIC | 4,455 | 1.01 (0.91,1.13) | 0.81 | 3,631 | 1.00 (0.92,1.09) | 0.97 |
|  | BHS | 638 | 1.04 (0.80,1.35) | 0.79 | 440 | 0.88 (0.66,1.18) | 0.39 |
|  | CHS | 1,574 | 0.91 (0.77,1.07) | 0.24 | 1,082 | 0.91 (0.76,1.10) | 0.33 |
|  | MESA | 2,022 | 0.94 (0.80,1.11) | 0.47 | 1,805 | 1.00 (0.85,1.18) | 0.97 |
|  | SOL | 4,276 | 1.03 (0.89,1.21) | 0.67 | 2,762 | 1.05 (0.92,1.20) | 0.45 |
| BMI adjusted OSA | ARIC | 4,455 | **1.23 (0.97,1.56)** | **0.09** | 3,631 | 1.02 (0.86,1.20) | 0.84 |
|  | BHS | 638 | 1.70 (0.64,4.54) | 0.29 | 440 | 0.88 (0.31,2.44) | 0.80 |
|  | CHS | 1,574 | 0.86 (0.55,1.35) | 0.51 | 1,082 | 1.06 (0.62,1.80) | 0.83 |
|  | MESA | 2,022 | 1.12 (0.77,1.62) | 0.57 | 1,805 | 1.23 (0.83,1.82) | 0.30 |
|  | SOL | 4,276 | 1.04 (0.79,1.37) | 0.80 | 2,762 | **1.22 (0.98,1.52)** | **0.08** |
| BMI unadjusted OSA | ARIC | 4,455 | 0.93 (0.81,1.06) | 0.29 | 3,631 | 1.00 (0.90,1.10) | 0.93 |
|  | BHS | 638 | 1.39 (0.69,2.82) | 0.36 | 440 | 0.66 (0.31,1.42) | 0.29 |
|  | CHS | 1,574 | 1.10 (0.77,1.58) | 0.60 | 1,082 | 0.92 (0.61,1.39) | 0.69 |
|  | MESA | 2,022 | 1.15 (0.84,1.57) | 0.39 | 1,805 | 1.03 (0.74,1.42) | 0.87 |
|  | SOL | 4,276 | 1.07 (0.85,1.35) | 0.57 | 2,762 | 1.13 (0.91,1.39) | 0.27 |
| **Sleep PGS** | **Cohort** | **GCF Change** | | | | | |
|  |  | **Females** | | | **Males** | | |
|  |  | **N** | **Estimate** | ***P*** | **N** | **Estimate** | ***P*** |
|  |  |  | **(95% CI)** |  |  | **(95% CI)** |  |
| Insomnia | ARIC | 2,029 | 0.01 (-0.04,0.05) | 0.74 | 1,501 | -0.03 (-0.07,0.02) | 0.21 |
|  | BHS | 360 | -0.01 (-0.22,0.21) | 0.95 | 225 | -0.22 (-0.48,0.04) | 0.10 |
|  | CHS | 997 | **-0.05 (-0.10,0.00)** | **0.07** | 642 | -0.04 (-0.09,0.02) | 0.16 |
|  | MESA | 788 | -0.03 (-0.09,0.03) | 0.28 | 715 | 0.02 (-0.04,0.07) | 0.53 |
|  | SOL | 2,938 | 0.01 (-0.03,0.05) | 0.77 | 1,701 | 0.03 (-0.01,0.08) | 0.16 |
| Long Sleep | ARIC | 2,029 | -0.01 (-0.05,0.03) | 0.77 | 1,501 | 0.00 (-0.05,0.04) | 0.90 |
|  | BHS | 360 | 0.05 (-0.12,0.23) | 0.55 | 225 | 0.18 (-0.02,0.37) | 0.08 |
|  | CHS | 997 | 0.00 (-0.04,0.04) | 0.86 | 642 | -0.03 (-0.08,0.02) | 0.20 |
|  | MESA | 788 | -0.01 (-0.05,0.04) | 0.76 | 715 | -0.02 (-0.06,0.02) | 0.26 |
|  | SOL | 2,938 | 0.00 (-0.04,0.03) | 0.86 | 1,701 | 0.02 (-0.05,0.10) | 0.53 |
| Short Sleep | ARIC | 2,029 | -0.01 (-0.05,0.03) | 0.68 | 1,501 | -0.03 (-0.07,0.02) | 0.21 |
|  | BHS | 360 | 0.11 (-0.13,0.35) | 0.36 | 225 | 0.00 (-0.30,0.29) | 0.98 |
|  | CHS | 997 | **-0.05 (-0.10,0.01)** | **0.09** | 642 | -0.02 (-0.08,0.04) | 0.53 |
|  | MESA | 788 | 0.00 (-0.07,0.06) | 0.90 | 715 | 0.01 (-0.05,0.07) | 0.71 |
|  | SOL | 2,938 | 0.03 (-0.01,0.07) | 0.16 | 1,701 | -0.02 (-0.07,0.02) | 0.34 |
| Sleep Duration | ARIC | 2,029 | 0.01 (-0.03,0.06) | 0.57 | 1,501 | 0.00 (-0.04,0.05) | 0.84 |
|  | BHS | 360 | 0.00 (-0.18,0.18) | 1.00 | 225 | 0.19 (-0.04,0.43) | 0.11 |
|  | CHS | 997 | **0.06 (0.01,0.10)** | **0.01** | 642 | -0.01 (-0.06,0.04) | 0.62 |
|  | MESA | 788 | 0.01 (-0.04,0.06) | 0.66 | 715 | **-0.04 (-0.09,0.01)** | **0.13** |
|  | SOL | 2,938 | -0.03 (-0.07,0.02) | 0.23 | 1,701 | 0.03 (-0.03,0.10) | 0.35 |
| Daytime Sleepiness | ARIC | 2,029 | 0.01 (-0.03,0.05) | 0.61 | 1,501 | 0.00 (-0.05,0.04) | 0.92 |
|  | BHS | 360 | 0.01 (-0.14,0.16) | 0.91 | 225 | -0.05 (-0.25,0.15) | 0.63 |
|  | CHS | 997 | 0.01 (-0.03,0.05) | 0.66 | 642 | -0.01 (-0.05,0.04) | 0.79 |
|  | MESA | 788 | -0.03 (-0.08,0.02) | 0.26 | 715 | 0.00 (-0.05,0.04) | 0.82 |
|  | SOL | 2,938 | 0.01 (-0.03,0.05) | 0.57 | 1,701 | 0.00 (-0.04,0.05) | 0.94 |
| BMI adjusted OSA | ARIC | 2,029 | -0.03 (-0.12,0.05) | 0.48 | 1,501 | -0.02 (-0.10,0.06) | 0.69 |
|  | BHS | 360 | 0.05 (-0.52,0.62) | 0.86 | 225 | 0.17 (-0.56,0.91) | 0.64 |
|  | CHS | 997 | -0.01 (-0.13,0.10) | 0.83 | 642 | 0.01 (-0.12,0.15) | 0.83 |
|  | MESA | 788 | 0.02 (-0.09, 0.14) | 0.69 | 715 | 0.01 (-0.10,0.12) | 0.85 |
|  | SOL | 2,938 | -0.05 (-0.12,0.03) | 0.22 | 1,701 | **0.14 (0.03,0.25)** | **0.01** |
| BMI unadjusted OSA | ARIC | 2,029 | 0.01 (-0.04, 0.06) | 0.66 | 1,501 | 0.02 (-0.03,0.06) | 0.52 |
|  | BHS | 360 | 0.11 (-0.31, 0.53) | 0.60 | 225 | -0.20 (-0.76,0.36) | 0.48 |
|  | CHS | 997 | -0.04 (-0.13, 0.05) | 0.35 | 642 | 0.00 (-0.10,0.10) | 0.99 |
|  | MESA | 788 | -0.01 (-0.11, 0.08) | 0.80 | 715 | 0.01 (-0.07,0.10) | 0.79 |
|  | SOL | 2,938 | 0.00 (-0.07, 0.06) | 0.93 | 1,701 | 0.05 (-0.04,0.13) | 0.27 |
| **Sleep PGS** | **Cohort** | **Incident Poor GCF** | | | | | |
|  |  | **Females** | | | **Males** | | |
|  |  | **N** | **IRR** | ***P*** | **N** | **IRR** | ***P*** |
|  |  |  | **(95% CI)** |  |  | **(95% CI)** |  |
| Insomnia | ARIC | 1,921 | 1.08 (0.93,1.26) | 0.32 | 1,275 | 0.91 (0.79,1.06) | 0.24 |
|  | BHS | 309 | **0.70 (0.55,0.88)** | **<0.01** | 192 | 0.88 (0.59,1.31) | 0.52 |
|  | CHS | 876 | 1.05 (0.83,1.34) | 0.67 | 559 | 0.80 (0.56,1.15) | 0.22 |
|  | MESA | 705 | 1.10 (0.80,1.52) | 0.56 | 647 | 0.94 (0.65,1.35) | 0.73 |
|  | SOL | 2,608 | 0.93 (0.77,1.12) | 0.43 | 1,359 | 1.03 (0.79,1.33) | 0.83 |
| Long Sleep | ARIC | 1,921 | 1.10 (0.96,1.27) | 0.18 | 1,275 | 0.99 (0.86,1.14) | 0.86 |
|  | BHS | 309 | **0.76 (0.60,0.95)** | **0.02** | 192 | 0.92 (0.63,1.32) | 0.64 |
|  | CHS | 876 | 1.05 (0.87,1.27) | 0.59 | 559 | 1.01 (0.75,1.37) | 0.93 |
|  | MESA | 705 | 1.00 (0.78,1.28) | 0.97 | 647 | 1.15 (0.88,1.50) | 0.32 |
|  | SOL | 2,608 | 1.13 (0.94,1.35) | 0.19 | 1,359 | 1.18 (0.86,1.61) | 0.31 |
| Short Sleep | ARIC | 1,921 | 0.99 (0.84,1.16) | 0.89 | 1,275 | 1.07 (0.92,1.25) | 0.35 |
|  | BHS | 309 | **0.60 (0.47,0.77)** | **<0.01** | 192 | **0.63 (0.40,0.98)** | **0.04** |
|  | CHS | 876 | 1.10 (0.87,1.41) | 0.43 | 559 | 0.93 (0.65,1.33) | 0.69 |
|  | MESA | 705 | 1.05 (0.74,1.49) | 0.77 | 647 | 0.87 (0.60,1.25) | 0.45 |
|  | SOL | 2,608 | 1.06 (0.92,1.23) | 0.42 | 1,359 | 1.07 (0.86,1.32) | 0.55 |
| Sleep Duration | ARIC | 1,921 | 0.94 (0.79,1.12) | 0.49 | 1,275 | 1.04 (0.90,1.21) | 0.57 |
|  | BHS | 309 | **1.44 (1.15,1.80)** | **<0.01** | 192 | 1.25 (0.85,1.85) | 0.26 |
|  | CHS | 876 | 0.93 (0.76,1.13) | 0.45 | 559 | 1.14 (0.83,1.56) | 0.42 |
|  | MESA | 705 | 0.88 (0.68,1.14) | 0.34 | 647 | 1.18 (0.87,1.59) | 0.28 |
|  | SOL | 2,608 | 1.03 (0.87,1.23) | 0.73 | 1,359 | 1.04 (0.78,1.40) | 0.78 |
| Daytime Sleepiness | ARIC | 1,921 | 0.92 (0.80,1.07) | 0.28 | 1,275 | 0.93 (0.81,1.08) | 0.34 |
|  | BHS | 309 | 0.96 (0.79,1.16) | 0.67 | 192 | 1.18 (0.81,1.72) | 0.40 |
|  | CHS | 876 | 0.93 (0.77,1.12) | 0.42 | 559 | 0.96 (0.72,1.30) | 0.80 |
|  | MESA | 705 | 1.19 (0.91,1.56) | 0.20 | 647 | 1.08 (0.81,1.45) | 0.58 |
|  | SOL | 2,608 | 0.95 (0.84,1.08) | 0.46 | 1,359 | 0.97 (0.81,1.16) | 0.72 |
| BMI adjusted OSA | ARIC | 1,921 | 1.24 (0.92,1.67) | 0.17 | 1,275 | **1.27 (0.97,1.67)** | **0.08** |
|  | BHS | 309 | **2.14 (1.49,3.08)** | **<0.01** | 192 | **2.61 (1.17,5.85)** | **0.02** |
|  | CHS | 876 | 0.80 (0.47,1.37) | 0.42 | 559 | 0.82 (0.35,1.96) | 0.66 |
|  | MESA | 705 | 1.16 (0.63,2.14) | 0.64 | 647 | 1.02 (0.52,1.98) | 0.96 |
|  | SOL | 2,608 | 0.87 (0.61,1.23) | 0.43 | 1,359 | 0.68 (0.44,1.05) | 0.08 |
| BMI unadjusted OSA | ARIC | 1,921 | 0.92 (0.77,1.10) | 0.37 | 1,275 | 1.08 (0.92,1.27) | 0.35 |
|  | BHS | 309 | **1.94 (1.41,2.67)** | **<0.01** | 192 | **2.37 (1.17,4.77)** | **0.02** |
|  | CHS | 876 | 1.11 (0.73,1.68) | 0.62 | 559 | 0.97 (0.52,1.81) | 0.92 |
|  | MESA | 705 | 1.51 (0.90,2.52) | 0.12 | 647 | 0.88 (0.50,1.55) | 0.66 |
|  | SOL | 2,608 | 1.08 (0.86,1.35) | 0.53 | 1,359 | 0.92 (0.69,1.23) | 0.57 |

**Table S10. Meta-analyzed effect estimates and heterogeneity test statistics for the associations of polygenic scores (PGS) for sleep traits and global cognitive outcomes in White, Black and Hispanic individuals.** Estimates are presented as differences in levels or odds ratio (OR) or incident risk ratio (IRR) and their 95% confidence interval (95% CI) per 1 standard deviation increase in each sleep trait PGS accounting for age, sex, race/ethnicity background, study locations, genetic PCs and education at baseline. For GCF change, the years interval between baseline and follow-up visit was additionally adjusted. For incident poor GCF, years interval between baseline and follow-up visit was fitted as an offset. Fixed-effects meta-analysis was applied using effect estimates from ARIC, BHS, CHS, MESA and SOL. Bolded entries indicate significant associations at z test p<0.05 or marginally significant associations at z test p<0.10.

| **Sleep PGS** | **Group** | **GCF** | | | | | **Poor GCF** | | | | |
| --- | --- | --- | --- | --- | --- | --- | --- | --- | --- | --- | --- |
|  |  | **N** | **Estimate**  **(95% CI)** | **Z test *P*** | **Q test *P*** | **I^2^** | **N** | **OR**  **(95% CI)** | **Z test *P*** | **Q test *P*** | **I^2^** |
| Insomnia | White | 11,119 | 0.00 (-0.02, 0.02) | 0.98 | 0.9 | 0 | 11,119 | 1.01 (0.94, 1.08) | 0.82 | 0.96 | 0 |
|  | Black | 3,174 | -0.01 (-0.05, 0.03) | 0.62 | 0.24 | 27.97 | 3,174 | 1.02 (0.90, 1.15) | 0.78 | 0.03 | 65.78* |
|  | Hispanic | 7,942 | 0.00 (-0.04, 0.03) | 0.80 | 0.57 | 0 | 7,942 | 0.99 (0.89, 1.10) | 0.87 | 0.29 | 12.33 |
| Long Sleep | White | 11,119 | **-0.02 (-0.04, 0.00)** | **0.03** | 0.89 | 0 | 11,119 | 1.04 (0.98, 1.11) | 0.17 | 0.27 | 24.15 |
|  | Black | 3,174 | 0.01 (-0.02, 0.04) | 0.48 | 0.26 | 24.41 | 3,174 | 0.98 (0.88, 1.08) | 0.64 | 0.07 | 57.79 |
|  | Hispanic | 7,942 | **-0.03 (-0.05, 0.00)** | **0.05** | 0.84 | 0 | 7,942 | **1.08 (0.99, 1.18)** | **0.08** | 0.62 | 0 |
| Short Sleep | White | 11,119 | 0.01 (-0.01, 0.03) | 0.15 | 0.04 | 64.35* | 11,119 | 0.96 (0.90, 1.02) | 0.16 | 0.23 | 31.12 |
|  | Black | 3,174 | -0.01 (-0.05, 0.03) | 0.51 | 0.29 | 19.21 | 3,174 | 1.03 (0.91, 1.16) | 0.68 | 0.9 | 0 |
|  | Hispanic | 7,942 | -0.01 (-0.03, 0.02) | 0.70 | 0.89 | 0 | 7,942 | 0.97 (0.88, 1.08) | 0.59 | 0.77 | 0 |
| Sleep Duration | White | 11,119 | -0.01 (-0.03, 0.01) | 0.18 | 0.22 | 31.52 | 11,119 | 1.03 (0.97, 1.09) | 0.38 | 0.49 | 0 |
|  | Black | 3,174 | 0.03 (-0.01, 0.06) | 0.13 | 0.53 | 0 | 3,174 | 0.97 (0.86, 1.08) | 0.54 | 0.54 | 0 |
|  | Hispanic | 7,942 | 0.00 (-0.03, 0.03) | 0.96 | 0.79 | 0 | 7,942 | 1.05 (0.96, 1.15) | 0.29 | 0.53 | 0 |
| Daytime Sleepiness | White | 11,119 | 0.00 (-0.02, 0.02) | 0.86 | 0.92 | 0 | 11,119 | 1.00 (0.94, 1.06) | 0.99 | 0.64 | 0 |
|  | Black | 3,174 | 0.01 (-0.02, 0.05) | 0.47 | 0.64 | 0 | 3,174 | 0.92 (0.82, 1.02) | 0.12 | 0.77 | 0 |
|  | Hispanic | 7,942 | -0.01 (-0.03, 0.02) | 0.63 | 0.11 | 60.18 | 7,942 | 1.03 (0.95, 1.12) | 0.50 | 0.58 | 0 |
| BMI adjusted OSA | White | 11,119 | -0.01 (-0.03, 0.02) | 0.62 | 0.26 | 24.62 | 11,119 | 1.04 (0.97, 1.11) | 0.30 | 0.5 | 0 |
|  | Black | 3,174 | 0.04 (-0.02, 0.09) | 0.21 | 0.12 | 48.14 | 3,174 | 0.96 (0.81, 1.14) | 0.64 | 0.42 | 0 |
|  | Hispanic | 7,942 | -0.05 (-0.10, 0.01) | 0.10 | 0.29 | 10.5 | 7,942 | **1.20 (1.02, 1.40)** | **0.03** | 0.09 | 64.75 |
| BMI unadjusted OSA | White | 11,119 | 0.01 (-0.01, 0.03) | 0.33 | 0.66 | 0 | 11,119 | 0.98 (0.91, 1.05) | 0.52 | 0.96 | 0 |
|  | Black | 3,174 | 0.04 (-0.02, 0.09) | 0.17 | 0.28 | 20.98 | 3,174 | 0.91 (0.78, 1.07) | 0.25 | 0.34 | 11.08 |
|  | Hispanic | 7,942 | **-0.05 (-0.10, 0.00)** | **0.03** | 0.7 | 0 | 7,942 | **1.14 (1.00, 1.32)** | **0.06** | 0.21 | 35.68 |
| **Sleep PGS** | **Group** | **GCF Change** | | | | | **Incident Poor GCF** | | | | |
|  |  | **N** | **Estimate**  **(95% CI)** | **Z test *P*** | **Q test *P*** | **I^2^** | **N** | **IRR**  **(95% CI)** | **Z test *P*** | **Q test *P*** | **I^2^** |
| Insomnia | White | 5,418 | **-0.02 (-0.04, 0.00)** | **0.05** | 0.17 | 39.56 | 4,977 | 0.99 (0.91, 1.08) | 0.84 | 0.76 | 0 |
|  | Black | 1,370 | -0.03 (-0.08, 0.02) | 0.22 | 0.55 | 0 | 1,143 | 0.99 (0.82, 1.20) | 0.90 | <0.01 | 85.56* |
|  | Hispanic | 4,925 | 0.02 (-0.01, 0.05) | 0.23 | 0.61 | 0 | 4,158 | 0.97 (0.83, 1.13) | 0.67 | 0.37 | 0 |
| Long Sleep | White | 5,418 | -0.01 (-0.03, 0.01) | 0.16 | 0.06 | 59.06 | 4,977 | **1.08 (0.99, 1.17)** | **0.08** | 0.17 | 40.04 |
|  | Black | 1,370 | 0.02 (-0.02, 0.06) | 0.28 | 0.61 | 0 | 1,143 | **0.85 (0.72, 1.01)** | **0.07** | 0.23 | 30.94 |
|  | Hispanic | 4,925 | 0.01 (-0.02, 0.05) | 0.52 | 0.93 | 0 | 4,158 | 1.06 (0.92, 1.22) | 0.41 | 0.32 | 0 |
| Short Sleep | White | 5,418 | -0.01 (-0.03, 0.01) | 0.31 | 0.65 | 0 | 4,977 | 1.01 (0.92, 1.10) | 0.89 | 0.56 | 0 |
|  | Black | 1,370 | **-0.06 (-0.11, -0.01)** | **0.03** | 0.98 | 0 | 1,143 | 0.99 (0.82, 1.20) | 0.94 | <0.01 | 76.63* |
|  | Hispanic | 4,925 | 0.00 (-0.03, 0.03) | 0.85 | 0.98 | 0 | 4,158 | 1.04 (0.91, 1.18) | 0.57 | 0.36 | 0 |
| Sleep Duration | White | 5,418 | 0.01 (-0.02, 0.03) | 0.61 | 0.37 | 5.15 | 4,977 | 1.04 (0.95, 1.13) | 0.39 | 0.81 | 0 |
|  | Black | 1,370 | **0.04 (0.00, 0.09)** | **0.06** | 0.87 | 0 | 1,143 | 0.96 (0.81, 1.14) | 0.65 | <0.01 | 82.40* |
|  | Hispanic | 4,925 | 0.00 (-0.03, 0.04) | 0.83 | 0.82 | 0 | 4,158 | 1.03 (0.89, 1.18) | 0.72 | 0.93 | 0 |
| Daytime Sleepiness | White | 5,418 | 0.00 (-0.02, 0.02) | 0.89 | 0.89 | 0 | 4,977 | 0.99 (0.91, 1.07) | 0.79 | 0.22 | 32.25 |
|  | Black | 1,370 | 0.03 (-0.02, 0.07) | 0.22 | 0.58 | 0 | 1,143 | 0.87 (0.73, 1.03) | 0.11 | 0.86 | 0 |
|  | Hispanic | 4,925 | -0.01 (-0.04, 0.02) | 0.67 | 0.02 | 80.51* | 4,158 | 0.99 (0.89, 1.11) | 0.88 | 0.15 | 50.91 |
| BMI adjusted OSA | White | 5,418 | -0.01 (-0.04, 0.02) | 0.49 | 0.89 | 0 | 4,977 | **1.12 (1.01, 1.25)** | **0.03** | 0.27 | 24 |
|  | Black | 1,370 | 0.01 (-0.06, 0.09) | 0.73 | 0.19 | 36.53 | 1,143 | 1.17 (0.90, 1.51) | 0.24 | 0.45 | 0 |
|  | Hispanic | 4,925 | 0.05 (-0.01, 0.11) | 0.12 | 0.43 | 0 | 4,158 | 0.85 (0.64, 1.12) | 0.25 | 0.51 | 0 |
| BMI unadjusted OSA | White | 5,418 | 0.00 (-0.03, 0.03) | 1.00 | 0.11 | 49.69 | 4,977 | 1.00 (0.91, 1.12) | 0.93 | 0.18 | 38.42 |
|  | Black | 1,370 | 0.02 (-0.05, 0.09) | 0.63 | 0.3 | 17.45 | 1,143 | 1.15 (0.92, 1.45) | 0.22 | 0.65 | 0 |
|  | Hispanic | 4,925 | 0.04 (-0.01, 0.09) | 0.12 | 0.06 | 72.41 | 4,158 | 1.04 (0.85, 1.26) | 0.72 | 0.14 | 54.91 |

*I-squared is statistically significant (at Q test p<0.05), indicating potential effect heterogeneity across the meta-analysis cohorts.

**Table S11. Race/ethnicity specific associations of polygenic scores (PGS) for sleep traits and global cognitive outcomes in each cohort.** Estimates are presented as differences in levels or odds ratio (OR) or incident risk ratio (IRR) and their 95% confidence interval (95% CI) per 1 standard deviation increase in each sleep trait PGS accounting for age, sex, study locations, genetic PCs and education at baseline. For GCF change, the years interval between baseline and follow-up visit was additionally adjusted. For incident poor GCF, years interval between baseline and follow-up visit was fitted as an offset. Bolded entries indicate significant associations at z test p<0.05 or marginally significant associations at z test p<0.10.

| **Sleep PGS** | **Cohort** | **GCF** | | | | | | | | |
| --- | --- | --- | --- | --- | --- | --- | --- | --- | --- | --- |
|  |  | **White** | | | **Black** | | | **Hispanic** | | |
|  |  | **N** | **Estimate (95% CI)** | ***P*** | **N** | **Estimate (95% CI)** | ***P*** | **N** | **Estimate (95% CI)** | ***P*** |
| Insomnia | ARIC | 6,688 | 0.00 (-0.03,0.02) | 0.85 | 1,398 | 0.03 (-0.03,0.08) | 0.34 | - | - | - |
|  | BHS | 715 | 0.01 (-0.07,0.09) | 0.78 | 363 | -0.08 (-0.21,0.04) | 0.20 | - | - | - |
|  | CHS | 2,143 | 0.01 (-0.03,0.05) | 0.60 | 501 | -0.07 (-0.17,0.04) | 0.20 | - | - | - |
|  | MESA | 1,573 | -0.01 (-0.05,0.03) | 0.65 | 912 | -0.02 (-0.09,0.05) | 0.60 | 904 | -0.02 (-0.09,0.05) | 0.54 |
|  | SOL | -* | - | - | - | - | - | 7,038 | 0.00 (-0.04,0.04) | 0.96 |
| Long Sleep | ARIC | 6,688 | **-0.02 (-0.05,0.00)** | **0.08** | 1,398 | **0.05 (0.00,0.11)** | **0.06** | - | - | - |
|  | BHS | 715 | -0.03 (-0.09,0.03) | 0.34 | 363 | 0.01 (-0.09,0.11) | 0.85 | - | - | - |
|  | CHS | 2,143 | -0.02 (-0.05,0.02) | 0.34 | 501 | 0.01 (-0.07,0.09) | 0.79 | - | - | - |
|  | MESA | 1,573 | -0.01 (-0.04,0.02) | 0.60 | 912 | -0.02 (-0.07,0.03) | 0.37 | 904 | -0.02 (-0.07,0.03) | 0.42 |
|  | SOL | - | - | - | - | - | - | 7,038 | **-0.03 (-0.06,0.00)** | **0.07** |
| Short Sleep | ARIC | 6,688 | 0.02 (-0.01,0.05) | 0.13 | 1,398 | 0.00 (-0.05,0.06) | 0.93 | - | - | - |
|  | BHS | 715 | **-0.10 (-0.19,-0.01)** | **0.03** | 363 | 0.06 (-0.09,0.21) | 0.44 | - | - | - |
|  | CHS | 2,143 | **0.04 (0.00,0.08)** | **0.08** | 501 | **-0.10 (-0.21,0.01)** | **0.07** | - | - | - |
|  | MESA | 1,573 | 0.00 (-0.05,0.04) | 0.90 | 912 | -0.02 (-0.09,0.05) | 0.59 | 904 | 0.00 (-0.08,0.07) | 0.99 |
|  | SOL | - | - | - | - | - | - | 7,038 | -0.01 (-0.04,0.02) | 0.68 |
| Sleep Duration | ARIC | 6,688 | -0.02 (-0.04,0.01) | 0.25 | 1,398 | 0.03 (-0.02,0.09) | 0.23 | - | - | - |
|  | BHS | 715 | 0.04 (-0.03,0.11) | 0.26 | 363 | -0.06 (-0.19,0.07) | 0.35 | - | - | - |
|  | CHS | 2,143 | **-0.03 (-0.07,0.00)** | **0.06** | 501 | 0.02 (-0.07,0.11) | 0.70 | - | - | - |
|  | MESA | 1,573 | 0.00 (-0.03,0.04) | 0.89 | 912 | 0.04 (-0.02,0.10) | 0.16 | 904 | 0.01 (-0.05,0.06) | 0.84 |
|  | SOL | - | - | - | - | - | - | 7,038 | 0.00 (-0.03,0.03) | 0.86 |
| Daytime Sleepiness | ARIC | 6,688 | 0.00 (-0.03,0.02) | 0.75 | 1,398 | 0.02 (-0.04,0.07) | 0.51 | - | - | - |
|  | BHS | 715 | 0.00 (-0.05,0.06) | 0.88 | 363 | 0.00 (-0.11,0.10) | 0.97 | - | - | - |
|  | CHS | 2,143 | -0.01 (-0.04,0.02) | 0.65 | 501 | 0.05 (-0.03,0.14) | 0.20 | - | - | - |
|  | MESA | 1,573 | 0.01 (-0.03,0.04) | 0.67 | 912 | -0.01 (-0.06,0.05) | 0.75 | 904 | 0.03 (-0.02,0.09) | 0.26 |
|  | SOL | - | - | - | - | - | - | 7,038 | -0.02 (-0.05,0.01) | 0.23 |
| BMI adjusted OSA | ARIC | 6,688 | -0.01 (-0.03,0.02) | 0.52 | 1,398 | **0.06 (0.00,0.13)** | **0.07** | - | - | - |
|  | BHS | 715 | -0.16 (-0.37,0.06) | 0.16 | 363 | -0.01 (-0.40,0.37) | 0.95 | - | - | - |
|  | CHS | 2,143 | 0.06 (-0.03,0.16) | 0.19 | 501 | **-0.23 (-0.46,0.00)** | **0.05** | - | - | - |
|  | MESA | 1,573 | -0.01 (-0.10,0.07) | 0.77 | 912 | 0.03 (-0.10,0.15) | 0.68 | 904 | **-0.11 (-0.23,0.02)** | **0.09** |
|  | SOL | - | - | - | - | - | - | 7,038 | -0.03 (-0.09,0.03) | 0.30 |
| BMI unadjusted OSA | ARIC | 6,688 | 0.02 (-0.01,0.04) | 0.23 | 1,398 | **0.06 (0.00,0.12)** | **0.07** | - | - | - |
|  | BHS | 715 | -0.07 (-0.23,0.09) | 0.38 | 363 | -0.02 (-0.30,0.26) | 0.90 | - | - | - |
|  | CHS | 2,143 | 0.02 (-0.06,0.09) | 0.64 | 501 | -0.13 (-0.31,0.05) | 0.16 | - | - | - |
|  | MESA | 1,573 | -0.01 (-0.08,0.06) | 0.76 | 912 | 0.03 (-0.07,0.14) | 0.53 | 904 | -0.07 (-0.17,0.03) | 0.19 |
|  | SOL | - | - | - | - | - | - | 7,038 | **-0.05 (-0.10,0.00)** | **0.08** |
| **Sleep PGS** | **Cohort** | **GCF Change** | | | | | | | | |
|  |  | **White** | | | **Black** | | | **Hispanic** | | |
|  |  | **N** | **Estimate (95% CI)** | ***P*** | **N** | **Estimate (95% CI)** | ***P*** | **N** | **Estimate (95% CI)** | ***P*** |
| Insomnia | ARIC | 3,008 | 0.00 (-0.03,0.03) | 0.98 | 522 | **-0.07 (-0.15,0.01)** | **0.10** | - | - | - |
|  | BHS | 425 | -0.13 (-0.31,0.06) | 0.18 | 160 | -0.06 (-0.42,0.30) | 0.74 | - | - | - |
|  | CHS | 1,339 | **-0.05 (-0.09,-0.01)** | **0.02** | 295 | -0.05 (-0.16,0.07) | 0.42 | - | - | - |
|  | MESA | 646 | -0.04 (-0.09,0.02) | 0.20 | 393 | 0.02 (-0.07,0.10) | 0.71 | 286 | -0.01 (-0.11,0.09) | 0.89 |
|  | SOL | - | - | - | - | - | - | 4,639 | **0.02 (-0.01,0.05)** | 0.19 |
| Long Sleep | ARIC | 3,008 | -0.01 (-0.04,0.02) | 0.66 | 522 | 0.02 (-0.06,0.10) | 0.66 | - | - | - |
|  | BHS | 425 | **0.13 (-0.01,0.27)** | **0.07** | 160 | 0.01 (-0.30,0.32) | 0.94 | - | - | - |
|  | CHS | 1,339 | -0.01 (-0.04,0.03) | 0.67 | 295 | -0.02 (-0.10,0.06) | 0.68 | - | - | - |
|  | MESA | 646 | **-0.05 (-0.09,-0.01)** | **0.02** | 393 | 0.05 (-0.01,0.12) | 0.11 | 286 | 0.01 (-0.06,0.09) | 0.69 |
|  | SOL | - | - | - | - | - | - | 4,639 | 0.01 (-0.03,0.05) | 0.61 |
| Short Sleep | ARIC | 3,008 | -0.01 (-0.04,0.02) | 0.41 | 522 | -0.05 (-0.13,0.03) | 0.21 | - | - | - |
|  | BHS | 425 | 0.09 (-0.12,0.29) | 0.42 | 160 | -0.07 (-0.46,0.32) | 0.72 | - | - | - |
|  | CHS | 1,339 | -0.02 (-0.07,0.02) | 0.26 | 295 | -0.08 (-0.19,0.03) | 0.16 | - | - | - |
|  | MESA | 646 | 0.01 (-0.05,0.06) | 0.78 | 393 | -0.05 (-0.15,0.04) | 0.25 | 286 | 0.00 (-0.10,0.11) | 0.98 |
|  | SOL | - | - | - | - | - | - | 4,639 | 0.00 (-0.03,0.03) | 0.85 |
| Sleep Duration | ARIC | 3,008 | 0.01 (-0.02,0.04) | 0.69 | 522 | 0.03 (-0.05,0.12) | 0.40 | - | - | - |
|  | BHS | 425 | 0.07 (-0.09,0.22) | 0.41 | 160 | 0.12 (-0.22,0.46) | 0.48 | - | - | - |
|  | CHS | 1,339 | 0.02 (-0.02,0.06) | 0.26 | 295 | 0.07 (-0.02,0.16) | 0.13 | - | - | - |
|  | MESA | 646 | -0.03 (-0.07,0.02) | 0.25 | 393 | 0.03 (-0.04,0.10) | 0.43 | 286 | 0.01 (-0.07,0.09) | 0.77 |
|  | SOL | - | - | - | - | - | - | 4,639 | 0.00 (-0.04,0.04) | 0.92 |
| Daytime Sleepiness | ARIC | 3,008 | 0.00 (-0.03,0.04) | 0.80 | 522 | 0.00 (-0.08,0.08) | 0.94 | - | - | - |
|  | BHS | 425 | -0.05 (-0.18,0.08) | 0.48 | 160 | 0.20 (-0.10,0.50) | 0.19 | - | - | - |
|  | CHS | 1,339 | 0.00 (-0.03,0.03) | 0.90 | 295 | 0.04 (-0.04,0.12) | 0.34 | - | - | - |
|  | MESA | 646 | -0.01 (-0.05,0.04) | 0.81 | 393 | 0.03 (-0.04,0.10) | 0.38 | 286 | **-0.10 (-0.18,-0.01)** | **0.02** |
|  | SOL | - | - | - | - | - | - | 4,639 | 0.01 (-0.02,0.04) | 0.68 |
| BMI adjusted OSA | ARIC | 3,008 | -0.01 (-0.04,0.02) | 0.64 | 522 | -0.01 (-0.11,0.09) | 0.86 | - | - | - |
|  | BHS | 425 | 0.11 (-0.38,0.60) | 0.66 | 160 | 0.13 (-0.95,1.21) | 0.81 | - | - | - |
|  | CHS | 1,339 | -0.04 (-0.13,0.06) | 0.42 | 295 | **0.25 (0.02,0.48)** | **0.03** | - | - | - |
|  | MESA | 646 | -0.01 (-0.12,0.10) | 0.90 | 393 | -0.04 (-0.20,0.12) | 0.61 | 286 | 0.11 (-0.06,0.28) | 0.20 |
|  | SOL | - | - | - | - | - | - | 4,639 | 0.04 (-0.03,0.10) | 0.23 |
| BMI unadjusted OSA | ARIC | 3,008 | 0.02 (-0.01,0.05) | 0.25 | 522 | 0.01 (-0.08,0.10) | 0.83 | - | - | - |
|  | BHS | 425 | -0.08 (-0.45,0.29) | 0.67 | 160 | 0.47 (-0.34,1.29) | 0.26 | - | - | - |
|  | CHS | 1,339 | -0.05 (-0.12,0.03) | 0.21 | 295 | 0.13 (-0.05,0.31) | 0.15 | - | - | - |
|  | MESA | 646 | **-0.08 (-0.16,0.01)** | **0.09** | 393 | -0.05 (-0.19,0.09) | 0.50 | 286 | **0.17 (0.03,0.31)** | **0.02** |
|  | SOL | - | - | - | - | - | - | 4,639 | 0.02 (-0.04,0.08) | 0.47 |
| **Sleep PGS** | **Cohort** | **Poor GCF** | | | | | | | | |
|  |  | **White** | | | **Black** | | | **Hispanic** | | |
|  |  | **N** | **OR (95% CI)** | ***P*** | **N** | **OR (95% CI)** | ***P*** | **N** | **OR (95% CI)** | ***P*** |
| Insomnia | ARIC | 6,688 | 1.01 (0.94,1.09) | 0.78 | 1,398 | **0.87 (0.74,1.02)** | **0.09** | - | - | - |
|  | BHS | 715 | 0.93 (0.64,1.36) | 0.72 | 363 | **1.39 (0.95,2.01)** | **0.09** | - | - | - |
|  | CHS | 2,143 | 1.02 (0.86,1.21) | 0.79 | 501 | 1.25 (0.91,1.70) | 0.17 | - | - | - |
|  | MESA | 1,573 | 0.95 (0.68,1.34) | 0.78 | 912 | 1.16 (0.88,1.54) | 0.29 | 904 | 0.89 (0.72,1.11) | 0.31 |
|  | SOL | - | - | - | - | - | - | 7,038 | 1.02 (0.91,1.15) | 0.72 |
| Long Sleep | ARIC | 6,688 | **1.07 (0.99,1.14)** | **0.09** | 1,398 | **0.84 (0.72,0.98)** | **0.02** | - | - | - |
|  | BHS | 715 | 1.21 (0.93,1.57) | 0.15 | 363 | 1.08 (0.79,1.46) | 0.64 | - | - | - |
|  | CHS | 2,143 | 0.96 (0.83,1.10) | 0.53 | 501 | 1.10 (0.87,1.38) | 0.43 | - | - | - |
|  | MESA | 1,573 | 0.92 (0.71,1.20) | 0.53 | 912 | 1.13 (0.91,1.41) | 0.26 | 904 | 1.04 (0.89,1.23) | 0.60 |
|  | SOL | - | - | - | - | - | - | 7,038 | **1.10 (0.99,1.21)** | **0.08** |
| Short Sleep | ARIC | 6,688 | **0.94 (0.88,1.01)** | **0.10** | 1,398 | 1.00 (0.86,1.17) | 0.99 | - | - | - |
|  | BHS | 715 | 1.24 (0.82,1.87) | 0.31 | 363 | 0.97 (0.64,1.49) | 0.90 | - | - | - |
|  | CHS | 2,143 | 1.05 (0.88,1.26) | 0.58 | 501 | 1.14 (0.82,1.59) | 0.43 | - | - | - |
|  | MESA | 1,573 | 0.76 (0.52,1.09) | 0.13 | 912 | 1.06 (0.78,1.44) | 0.72 | 904 | 0.94 (0.74,1.19) | 0.62 |
|  | SOL | - | - | - | - | - | - | 7,038 | 0.98 (0.87,1.10) | 0.72 |
| Sleep Duration | ARIC | 6,688 | 1.04 (0.97,1.12) | 0.24 | 1,398 | 0.93 (0.80,1.09) | 0.37 | - | - | - |
|  | BHS | 715 | 0.95 (0.70,1.29) | 0.75 | 363 | 1.06 (0.74,1.53) | 0.74 | - | - | - |
|  | CHS | 2,143 | 0.94 (0.81,1.10) | 0.45 | 501 | 1.13 (0.86,1.48) | 0.39 | - | - | - |
|  | MESA | 1,573 | 1.16 (0.88,1.53) | 0.28 | 912 | 0.89 (0.68,1.14) | 0.35 | 904 | 1.00 (0.83,1.20) | 0.99 |
|  | SOL | - | - | - | - | - | - | 7,038 | 1.07 (0.96,1.19) | 0.22 |
| Daytime Sleepiness | ARIC | 6,688 | 1.01 (0.94,1.09) | 0.70 | 1,398 | 0.93 (0.80,1.09) | 0.39 | - | - | - |
|  | BHS | 715 | 1.09 (0.85,1.41) | 0.49 | 363 | 0.90 (0.66,1.22) | 0.49 | - | - | - |
|  | CHS | 2,143 | 0.94 (0.82,1.08) | 0.36 | 501 | 0.82 (0.64,1.06) | 0.13 | - | - | - |
|  | MESA | 1,573 | 0.94 (0.72,1.22) | 0.63 | 912 | 0.98 (0.77,1.24) | 0.87 | 904 | 0.99 (0.83,1.17) | 0.88 |
|  | SOL | - | - | - | - | - | - | 7,038 | 1.04 (0.95,1.15) | 0.39 |
| BMI adjusted OSA | ARIC | 6,688 | 1.05 (0.97,1.12) | 0.23 | 1,398 | 0.95 (0.79,1.15) | 0.61 | - | - | - |
|  | BHS | 715 | 1.53 (0.59,3.94) | 0.38 | 363 | 0.79 (0.26,2.38) | 0.67 | - | - | - |
|  | CHS | 2,143 | 0.79 (0.53,1.19) | 0.26 | 501 | 1.63 (0.81,3.27) | 0.17 | - | - | - |
|  | MESA | 1,573 | 1.01 (0.52,1.95) | 0.99 | 912 | 0.80 (0.47,1.34) | 0.39 | 904 | **1.64 (1.10,2.44)** | **0.02** |
|  | SOL | - | - | - | - | - | - | 7,038 | 1.13 (0.95,1.34) | 0.18 |
| BMI unadjusted OSA | ARIC | 6,688 | 0.98 (0.91,1.06) | 0.63 | 1,398 | 0.89 (0.75,1.06) | 0.20 | - | - | - |
|  | BHS | 715 | 0.87 (0.44,1.72) | 0.69 | 363 | 1.06 (0.48,2.35) | 0.89 | - | - | - |
|  | CHS | 2,143 | 0.93 (0.68,1.27) | 0.64 | 501 | 1.43 (0.83,2.47) | 0.20 | - | - | - |
|  | MESA | 1,573 | 0.91 (0.53,1.58) | 0.75 | 912 | 0.77 (0.50,1.18) | 0.23 | 904 | **1.39 (0.99,1.93)** | **0.05** |
|  | SOL | - | - | - | - | - | - | 7,038 | 1.10 (0.94,1.28) | 0.24 |
| **Sleep PGS** | **Cohort** | **Incident Poor GCF** | | | | | | | | |
|  |  | **White** | | | **Black** | | | **Hispanic** | | |
|  |  | **N** | **IRR (95% CI)** | ***P*** | **N** | **IRR (95% CI)** | ***P*** | **N** | **IRR (95% CI)** | ***P*** |
| Insomnia | ARIC | 2,712 | 0.98 (0.87,1.09) | 0.69 | 484 | 1.18 (0.93,1.51) | 0.18 | - | - | - |
|  | BHS | 389 | 1.06 (0.83,1.34) | 0.64 | 112 | **0.15 (0.06,0.35)** | **<0.01** | - | - | - |
|  | CHS | 1,241 | 0.96 (0.77,1.18) | 0.68 | 192 | 1.10 (0.62,1.94) | 0.74 | - | - | - |
|  | MESA | 635 | 1.27 (0.71,2.30) | 0.42 | 355 | 0.85 (0.57,1.27) | 0.44 | 191 | 1.09 (0.80,1.48) | 0.58 |
|  | SOL | - | - |  | - | - | - | 3,967 | 0.93 (0.78,1.11) | 0.42 |
| Long Sleep | ARIC | 2,712 | 1.06 (0.95,1.19) | 0.27 | 484 | 0.92 (0.73,1.16) | 0.47 | - | - | - |
|  | BHS | 389 | 0.96 (0.78,1.17) | 0.66 | 112 | 0.96 (0.30,3.05) | 0.95 | - | - | - |
|  | CHS | 1,241 | 1.14 (0.95,1.36) | 0.15 | 192 | **0.59 (0.40,0.87)** | **0.01** | - | - | - |
|  | MESA | 635 | **1.60 (1.04,2.47)** | **0.03** | 355 | 0.95 (0.69,1.29) | 0.72 | 191 | 0.96 (0.76,1.22) | 0.77 |
|  | SOL | - | - |  | - | - | - | 3,967 | 1.12 (0.94,1.34) | 0.21 |
| Short Sleep | ARIC | 2,712 | 1.04 (0.93,1.16) | 0.54 | 484 | 1.00 (0.79,1.25) | 0.98 | - | - | - |
|  | BHS | 389 | 0.86 (0.65,1.13) | 0.27 | 112 | **0.16 (0.06,0.46)** | **<0.01** | - | - | - |
|  | CHS | 1,241 | 1.02 (0.82,1.28) | 0.83 | 192 | 1.16 (0.69,1.97) | 0.57 | - | - | - |
|  | MESA | 635 | 0.80 (0.42,1.53) | 0.51 | 355 | 1.24 (0.77,1.97) | 0.38 | 191 | 0.90 (0.65,1.25) | 0.54 |
|  | SOL | - | - |  | - | - | - | 3,967 | 1.07 (0.93,1.22) | 0.38 |
| Sleep Duration | ARIC | 2,712 | 1.01 (0.90,1.13) | 0.86 | 484 | 0.92 (0.73,1.16) | 0.49 | - | - | - |
|  | BHS | 389 | 1.10 (0.89,1.36) | 0.39 | 112 | **4.84 (2.1,11.15)** | **<0.01** | - | - | - |
|  | CHS | 1,241 | 1.05 (0.87,1.27) | 0.58 | 192 | **0.66 (0.42,1.05)** | **0.08** | - | - | - |
|  | MESA | 635 | 1.24 (0.74,2.07) | 0.42 | 355 | 0.99 (0.69,1.41) | 0.94 | 191 | 1.04 (0.81,1.33) | 0.78 |
|  | SOL | - | - |  | - | - | - | 3,967 | 1.02 (0.86,1.22) | 0.80 |
| Daytime Sleepiness | ARIC | 2,712 | 0.94 (0.84,1.05) | 0.29 | 484 | 0.88 (0.69,1.11) | 0.29 | - | - | - |
|  | BHS | 389 | 1.16 (0.97,1.38) | 0.11 | 112 | 0.53 (0.17,1.66) | 0.27 | - | - | - |
|  | CHS | 1,241 | 0.94 (0.79,1.12) | 0.47 | 192 | 0.89 (0.61,1.30) | 0.55 | - | - | - |
|  | MESA | 635 | 1.11 (0.72,1.71) | 0.63 | 355 | 0.87 (0.62,1.22) | 0.43 | 191 | 1.17 (0.91,1.51) | 0.23 |
|  | SOL | - | - |  | - | - | - | 3,967 | 0.95 (0.84,1.08) | 0.44 |
| BMI adjusted OSA | ARIC | 2,712 | **1.12 (1.01,1.25)** | **0.04** | 484 | 1.12 (0.84,1.48) | 0.45 | - | - | - |
|  | BHS | 389 | **1.69 (0.94,3.02)** | **0.08** | 112 | **3.55 (0.87,14.53)** | **0.08** | - | - | - |
|  | CHS | 1,241 | 0.79 (0.48,1.30) | 0.36 | 192 | 0.97 (0.30,3.18) | 0.96 | - | - | - |
|  | MESA | 635 | 1.47 (0.43,5.03) | 0.54 | 355 | 1.29 (0.56,2.95) | 0.55 | 191 | 0.73 (0.44,1.23) | 0.24 |
|  | SOL | - | - |  | - | - | - | 3,967 | 0.90 (0.65,1.25) | 0.54 |
| BMI unadjusted OSA | ARIC | 2,712 | 0.97 (0.87,1.08) | 0.59 | 484 | 1.15 (0.89,1.48) | 0.29 | - | - | - |
|  | BHS | 389 | 1.38 (0.86,2.20) | 0.18 | 112 | 3.55 (0.59,21.33) | 0.17 | - | - | - |
|  | CHS | 1,241 | 1.09 (0.74,1.60) | 0.67 | 192 | 1.01 (0.45,2.28) | 0.98 | - | - | - |
|  | MESA | 635 | 2.17 (0.85,5.57) | 0.11 | 355 | 1.12 (0.55,2.25) | 0.76 | 191 | 0.77 (0.49,1.19) | 0.24 |
|  | SOL | - | - |  | - | - | - | 3,967 | 1.11 (0.90,1.38) | 0.33 |

^*^ indicates missing entry.

**Figure S1. Derivation of global cognitive function (GCF) measures in the study.** PCA, principal component analysis; PC1, first principal component; GCF, global cognitive function.

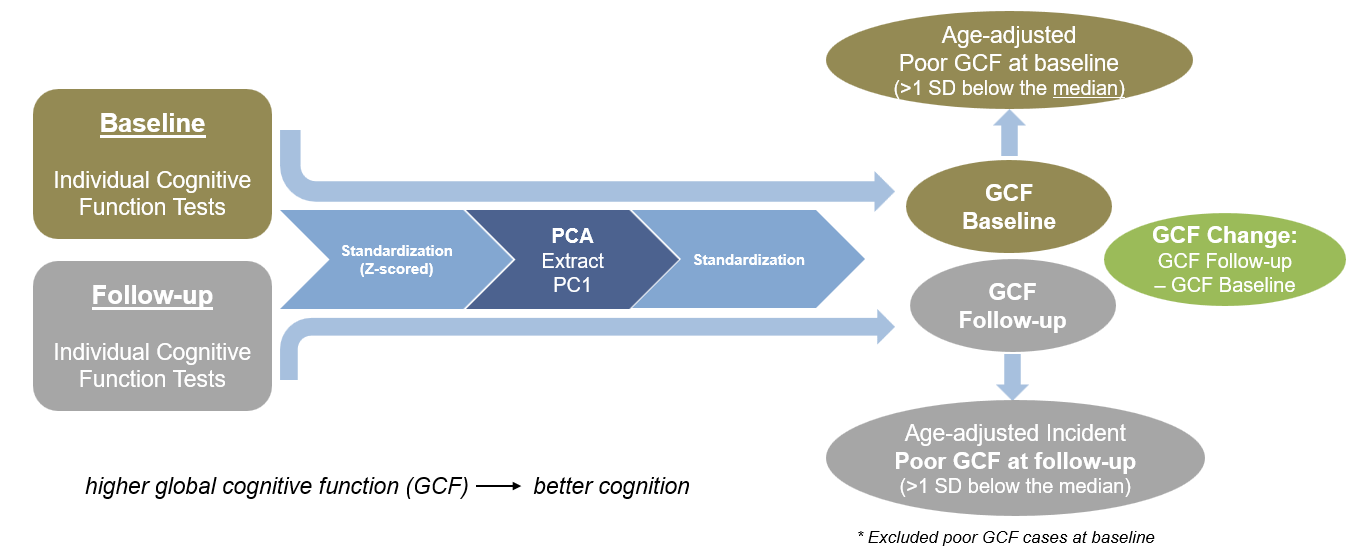
